# A Machine Learning Perspective on Causes of Suicides and identification of Vulnerable Categories using Multiple Algorithms

**DOI:** 10.1101/2021.04.08.21255162

**Authors:** Jitendra Shreemali, Prasun Chakrabarti, Tulika Chakrabarti, Sandeep Poddar, Daniel Sipple, Babak Kateb, Mohammad Nami

**Affiliations:** Professor, Techno India NJR Institute of Technology, Udaipur, Rajasthan, India; Provost, Techno India NJR Institute of Technology, Udaipur, Rajasthan, India; Sir Padampat Singhania University, Udaipur 313601, Rajasthan, India; Research and Innovation Vice Chancellery, Lincoln University College Malaysia; Midwest Spine and Brain Institute, Roseville, MN, and Society for Brain Mapping and Therapeutics, Los Angeles, CA, USA; Director, National Center for NanoBioElectoronics; Director, Brain Technology and Innovation Park; President, Brain Mapping Foundation; Chairman and CEO, Society for brain mapping and Therapeutics and, Chairman of Neuroscience20-G20 summit, Los Angeles, CA, USA; Neuroscience Center, Instituto de Investigaciones Científicas y Servicios de Alta Tecnología (INDICASAT AIP), City of Knowledge, Panama City, Panama; Department of Neuroscience, School of Advanced Medical Sciences and Technologies, Shiraz University of Medical Sciences; DANA Brain Health Institute, Iranian Neuroscience Society-Fars Chapter, Shiraz, Iran; Inclusive Brain Health, Swiss Alternative Medicine, Geneva, Switzerland

**Keywords:** Suicide, Happiness, Machine Learning, Neural Network, Dystopia

## Abstract

**Background:** Suicides represent a social tragedy with long term impact for the family. Given the growing incidence of suicides, a better understanding of factors causing it and addressing them has emerged as a social imperative.

**Material and Methods:** This study analyzed suicide data for three decades (1987-2016) and was carried out in two phases. Machine Learning Models run after pre-processing the suicide data included Neural network, Regression, Random Forest, XG Boost Tree, CHAID, Generalized Linear, Random Trees, Tree-AS and Auto Numeric Model.

**Results and Conclusion:** Analysis of findings suggested that the key predictors for suicide are Age, Gender, and Country. In the second phase, data from happiness reports were merged with suicide data to check if Country-specific factors impact the list or order of key predictors. While the key predictors remain the same, Country-specific factors like Generosity, Health and Trust impact the suicide rate in the Country.

## 1 Introduction

American Psychiatric Association defines suicide as self-inflicted death with evidence, implicit or explicit, showing that the person intended to die. A suicide attempt, on the other hand, is self-injurious behavior with a non-fatal outcome with evidence, implicit or explicit, of intent to die. Kahn (2019) defined suicide as the act of taking one’s life and quotes. Suicides are often measured in suicide rates that give a number of suicides per 100000 population. Rosston (2018) points to the 200% increase in suicide rates among those in the 15-24 age group in the period from the 1950s to mid-1990s with 443 suicides in 2016 by children in the age group of 9-14 years. The American Foundation for Suicide Prevention points out that suicide is the 10th leading cause of death in the USA, with the numbers dying on account of suicide being over 48000 in 2018 from an estimated 1.4 Million suicide attempts. In terms of sheer monetary value, suicide and self-inflicted injury cost the U.S. an estimated 69 Billion USD in 2015. Further, in 2018, the number of men dying on account of suicide was 3.56 times the number of women dying by suicide, with white males accounting for close to 70% of suicide deaths. Various factors have caused a continuous increase in the rate of suicides in the USA from 117.5/Million in 2009 to about 142/Million in 2018, an increase of over 20% in less than a decade. The rate of suicide differs across different age groups, with the highest rate in 2018 being seen among those in the 55-64 age group (202/Million), and the 45-54 age group is very close (200.4/Million). These figures in 2009 were 167 and 193, respectively, showing an increase in the Age of those dying on account of suicide. Another serious cause of concern is the jump in suicides for those in the 15-24 age group from 102/Million in 2009 to 144.5/Million in 2018, an increase of 41.67% in less than a decade with no instance of reduction in values in a subsequent year.

**Fig. 1:**
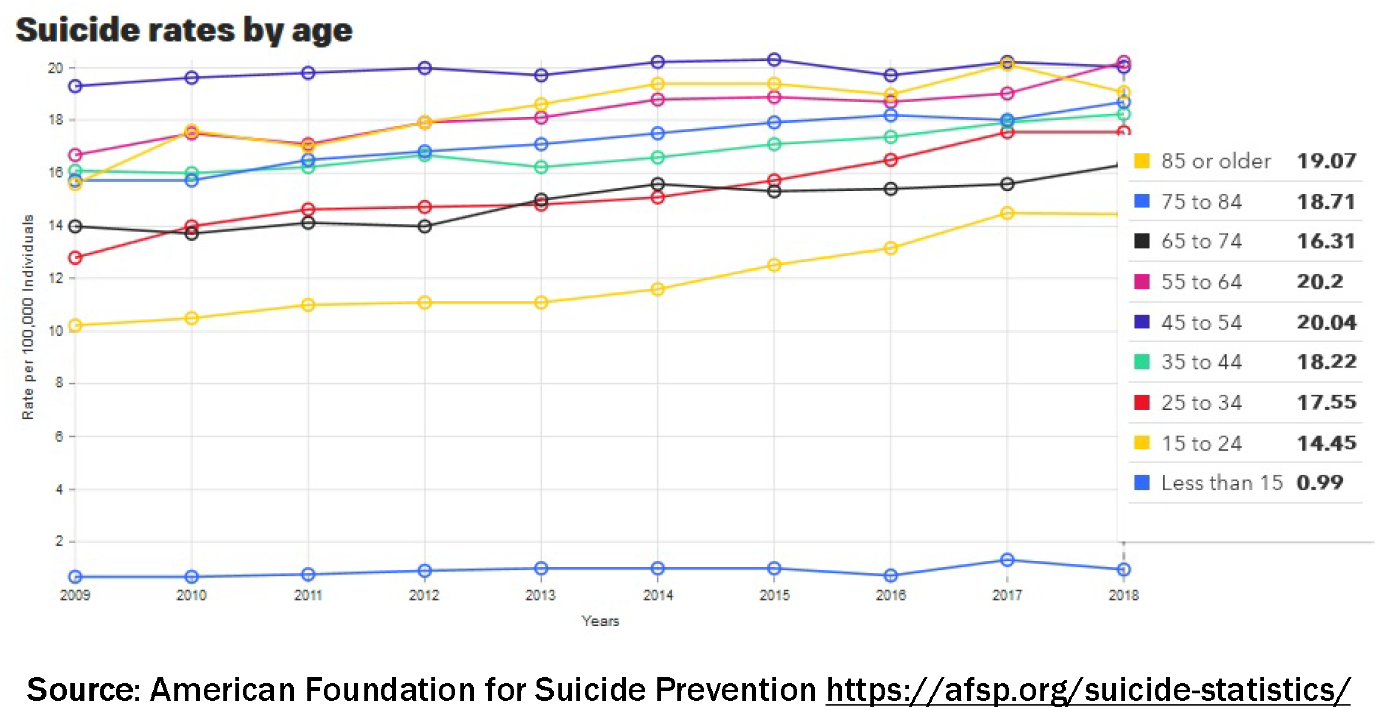
Breakdown of Suicide Rate by age in USA (2009-2018)

This increasing trend is also seen across the world, with 2014’s WHO report on preventing suicides stating that while suicides are preventable, the number of suicides across the world was close to 0.8 Million in the year 2012. It represented an annual age-standardized suicide rate of 114/Million (the rate for males being 150/Million and for females being 80/Million) and was the second leading cause of death among those in the 15-29 age-group. Further, it is the lower and middle-income countries witnessing about 75% of all suicides suggesting the role of income and means of livelihood as being possible factors in causing suicides.

**Fig. 2:**
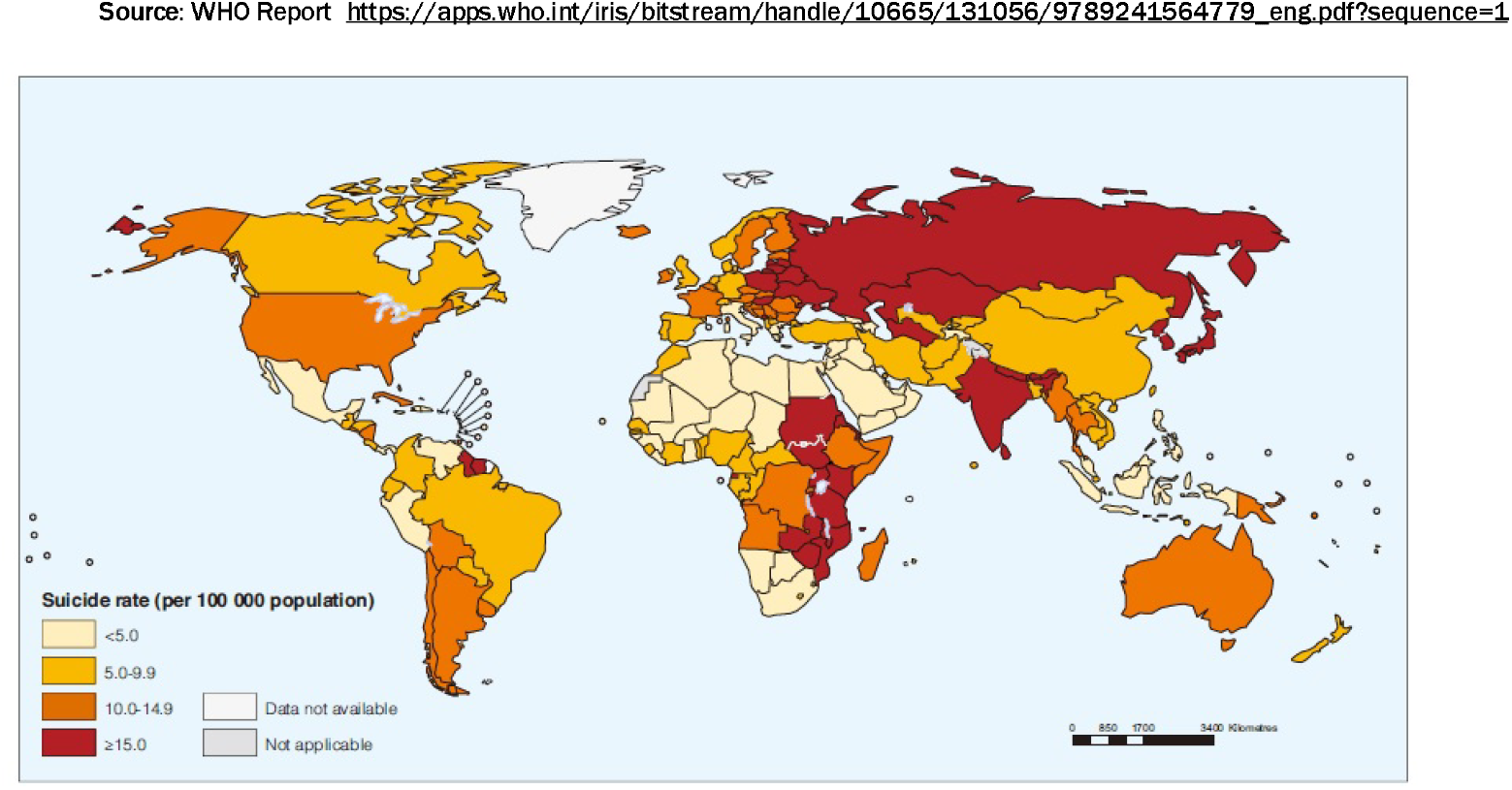
Suicide Rate/100000 population: Age Standardized for both Genders.

### LITERATURE REVIEW

As seen in the map above, suicides are a serious problem in all societies to different degrees, though its lower incidence in some parts of the world could provide clues on causes of suicides. One possible cause is the income group that the Country is in since higher income group countries provide more opportunities for progress. A review of data on total cases of suicides also suggests that the impact of on income is more pronounced on the female population with the Age-Standardized Suicide Rate/100000 population with suicide rate among females in all low as well as middle-income member states being at 87/Million as compared to 57 for the high-income member states. The larger percentage of total suicides from lower-and-middle-income countries, however, appears to be caused by a larger population living in these countries since higher income group countries account for 23.9% of global suicides despite the fact that their population is only 17.9% of the global population. This is further borne out by a relatively lower suicide rate in terms of standardized suicide rate (137/Million vs. 199/Million, i.e., over 45% higher rate in high-income countries).

**Fig. 3:**
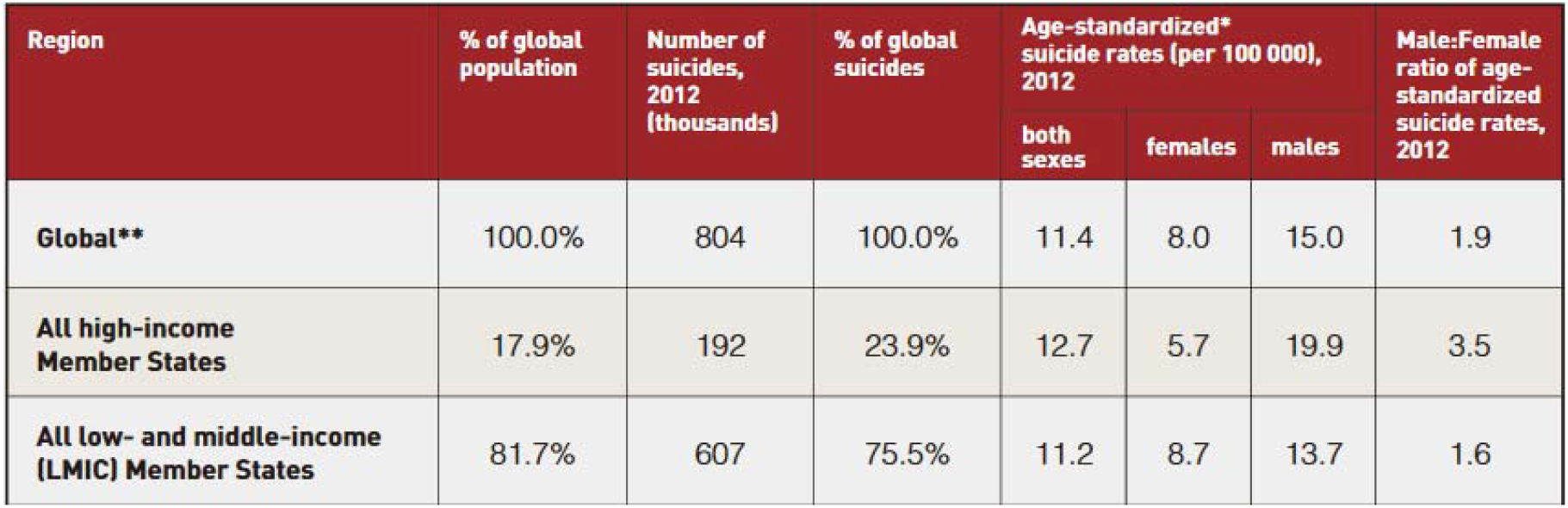
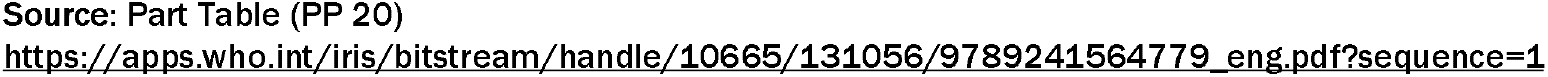
Number of Suicides in 2012 (estimated) based on Income Group of Member Country

Difference in suicides among different age groups in both categories of countries is shown below and again brings out the higher prevalence of suicides, as measured by suicide-rate, among higher income countries irrespective of age group.

**Fig. 4:**
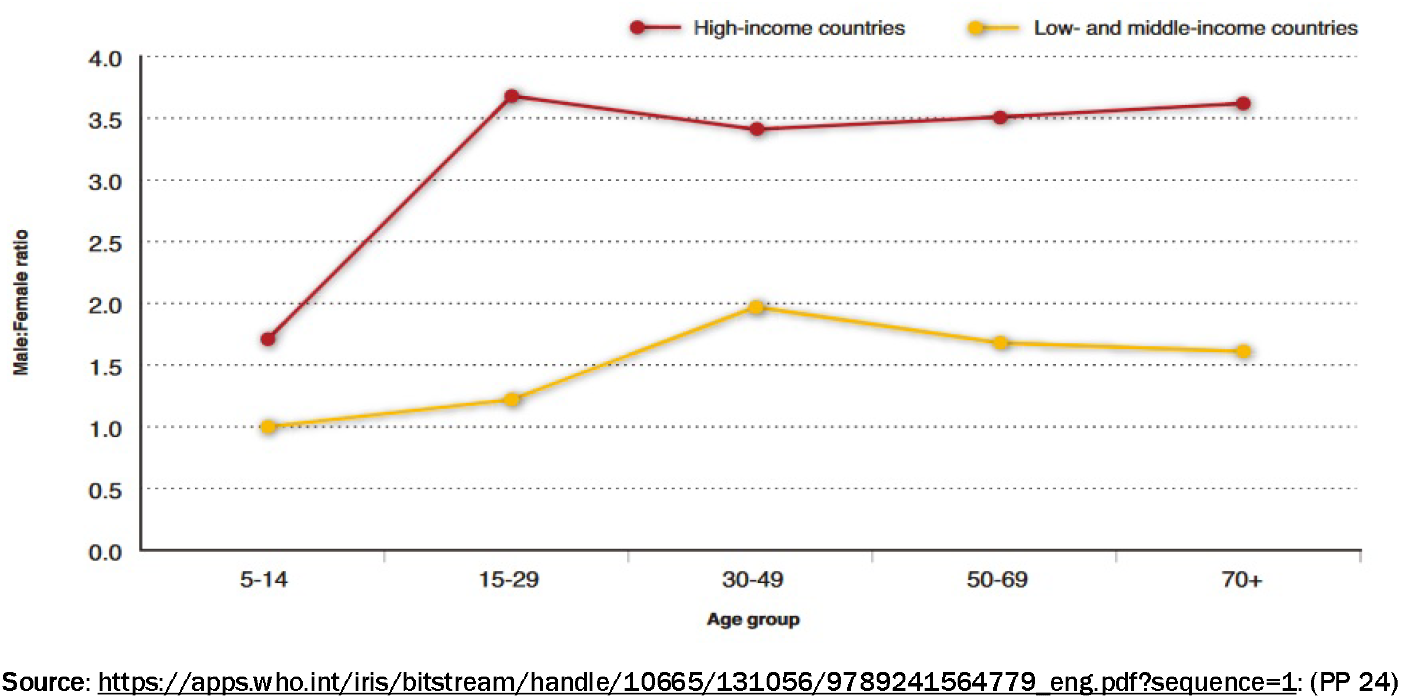
Ratio of Gender of Suicides (Male:Female Suicides) for Different Income-Group Counties

The Suicide Prevention Resource Centre views risk factors for suicide as falling at multiple levels: (i) Individual-level including personality traits, mental disorders, and genetic predisposition, and those beyond the individual. These include the community and family with risk factors including the degree of cohesion in the family, cohesive/dysfunctional relationships in the family, and availability of mental health and support services in the community. Other important risk factors include earlier suicide attempts, substance abuse, mood disorders, and availability of means for suicide.

Bilsen (2018) refers to suicide as being a leading cause of death in late childhood and adolescence with a disruptive psychosocial and adverse socio-economic impact. The key risk factors that contribute to suicidal behavior among the young include specific personality predisposition or mental disorders caused by genetic loading or family processes with possible triggering by psychosocial stressors coupled with available means for committing suicide and exposure to inspiring models. Bilsen’s review suggests ambivalence among those showing an inclination for suicide with the fatal transition happening on impulse; as a result, acute psychosocial stressor and availability of means to commit suicide. Kahn points to the possibility of gauging suicidal thoughts or tendencies by observing overt behavior that includes: (i) Threats or comments about suicide (e.g., saying that they have no reason to continue living, talking of being trapped or hopeless, or talking of suicide as a way out); (ii) Withdrawing from family and friends; (iii) Demonstrating reckless or unusually aggressive behavior accompanied with mood swings; (iv) Enhanced consumption of alcohol or stronger intoxicants. (v) Acquiring means doing personal harm (e.g., purchasing a gun). Suicide, being the second leading cause by the young, has been researched by WHO, and the 2016 data is reported below.

**Fig. 5:**
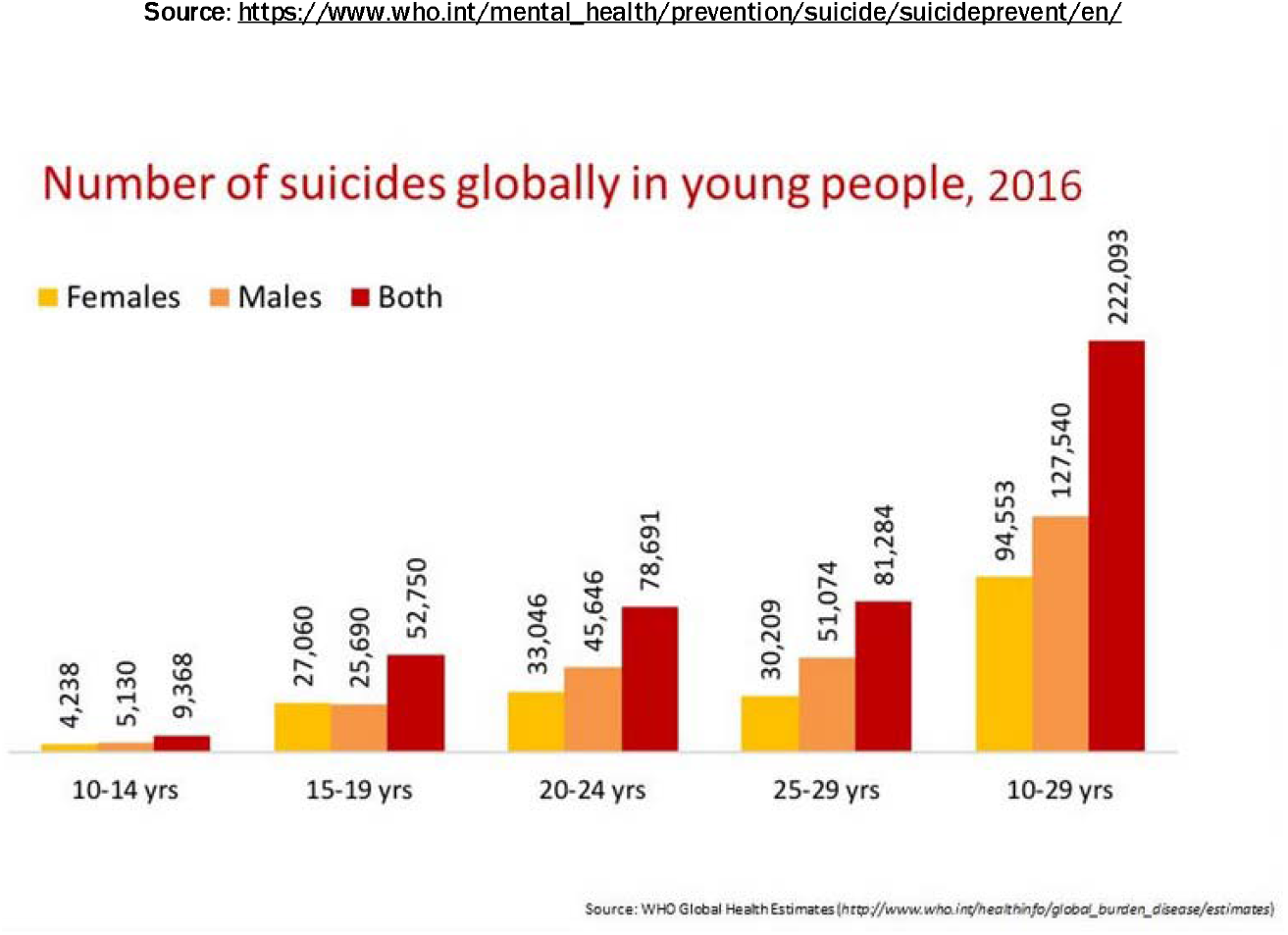
World Health Organization data on suicides by the young

Given the prevalence of suicide across all societies, suicide prevention continues to be a challenge across the world. However, an understanding of its causes and ways of giving effect to suicidal tendencies would surely help, as shown by the success of the intervention in the USA in 2010. The figure below highlights comparative successes among three approaches in preventing suicides. Removal of firearms from the individual, the most frequently used means for suicide, was seen to be almost 1.5 times as effective as psychotherapy in emergency care. In some cases, separating means of suicide can be more difficult, as in the case of suicide caused by jumping from bridges, high-rise buildings, or in front of trains since it would require constant watch and care.

**Fig. 6:**
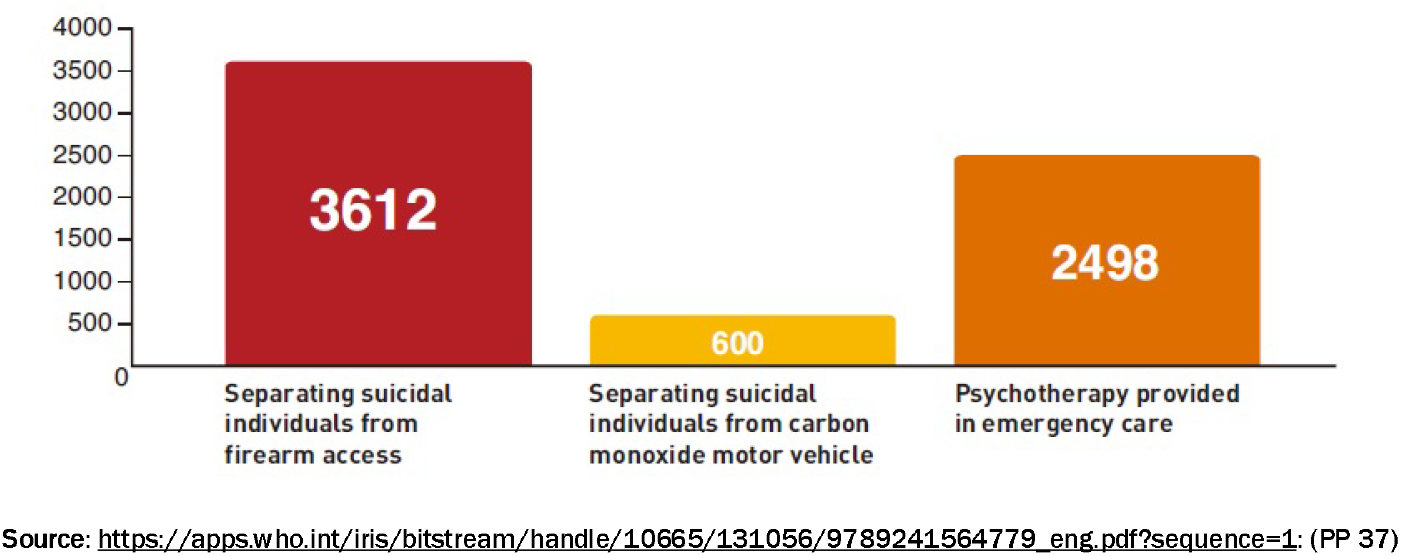
Preventing Suicide Deaths: Taking away means of suicide.

As per Vijaykumar (2010), India’s suicide rate was 10.3, having increased by 43% over the thirty years preceding it though the male to female suicide ratio in India remained at 1.4:1 and 71% suicides were by those less than 44 years of age. The ratio remaining the same would indicate a roughly identical increase in the percentage of suicides among males and females. Sathyavathi (1971) reported that in psychiatric patients, 64% of suicide attempts were from schizophrenia patients and close to 42% attempted suicide more than once often (about67%) employing the same method for attempting suicide. Besides schizophrenia, depression was the next important factor leading to suicides among psychiatric inpatients. Snowdon (2019) reported the suicide rate as increasing from 6.3 in 1978 to 8.9 in 1990 and then rising to 11.2 in 2011, with the ratio of male to females being 2.2. The male-to-female ratio happens to be much higher in Europe at 4:1 and close to 3.1:1 for six highly developed English-speaking countries, namely, USA, Canada, Australia, New-Zealand, Ireland, and the U.K. As far as the age of suicides goes, male suicides were seen to be bimodal, with one peak at 30-44 years and another after 70 years of age, while female suicides showed a peak in the 15-29 age group. Snowdon also reported a significant increase in the suicide rate among those more than 75 years old.

Icaza and Gorn (2016) observed, in their study of suicidal behavior, that while ideation occurs more in the very young in Mexico, the actual attempt is much less among the young, suggesting that more effort is required to wean the older generation away from suicide after ideation as compared to the young.

**Fig. 7:**
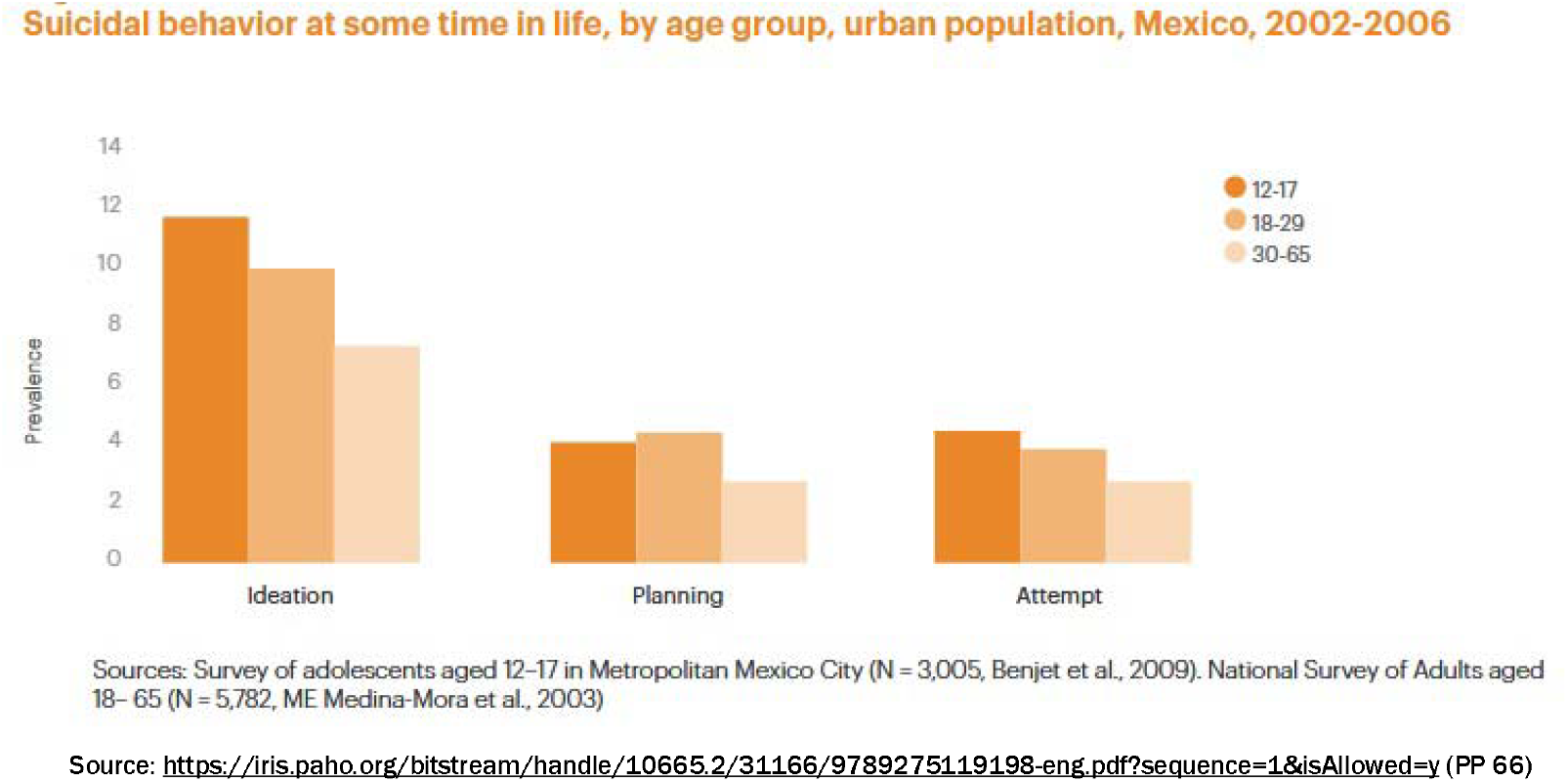
Stages of Suicidal Behaviour by Age Groups.

Picture This observes the prevalence of significant psychiatric illness among 60-90% of suicide victims when attempting suicide, the illness that may not necessarily have been diagnosed or treated. Two of the frequently observed psychiatric illness are mood disorders (depression, bipolar disorder or borderline personality disorder) and substance abuse. The combined prevalence of mood disorders and substance abuse greatly increases the risk of suicide. Similarly, depressed individuals showing open aggression, anxiety, or agitation could indicate a greatly enhanced risk of suicide.

The World Health Organization (2008) report highlight is that suicide prevention, though possible, presents big challenges because of the range of activities that range from conditions of bringing up children to effective treatment of psychiatric disorders as well as addressing environmental risk factors. Media reporting on suicides needs to be done carefully to reduce the possibility of imitative suicides, especially if a celebrity committed suicide with bit fan following. The extended gestation period for suicide prevention methods is also observed when Acevedo (2016) provides details of measures in Cuba that contributed to lowering the adjusted suicide rate from 23/100000 population in 1982 to 9.1/100000 in 2013. A significant role here was played by the National Program for Prevention of Suicidal Behavior developed in 1988 and is updated every five years based on the needs of the Country’s population. Its features include: (a) Training health, mental health workers, and community in aspects of mental health; (b) Setting up a health surveillance system aimed at studying every suicide as well as attempted suicide and provide (c) Care to every individual who attempted suicide.

Mann et al. (2005) find the presence of psychiatric disorders in at least 90% of suicides, with over 80% being under treatment. The study notes that depression is either not treated or inadequately treated even after attempting to commit suicide, leaving the root cause unaddressed. Treating psychiatric and mood disorders should be at the core of suicide prevention efforts. This is further supported by a striking correlation between the use of anti-depressants and the rate of untreated depression as well as suicides. Shaffer, Garland, Gould, Fisher, and Trautman (1998) state that as far as teenagers are concerned, most suicides occur among teenagers with the identifiable character or mental disorders. The National Action Programme for Suicide Prevention (2016) by The Public Health Agency of Sweden lists promoting good life opportunities for the less privileged (those with a low income or low educational level), reducing alcohol consumption (as alcohol reduces the individual’s ability to refrain from impulsive or risky actions) and reducing access to means of suicide as the three most important measures for suicide prevention.

### METHODOLOGY

Data for the study was taken from the publicly available Kaggle sites, namely, https://www.kaggle.com/russellyates88/suicide-rates-overview-1985-to-2016 for suicide data and https://www.kaggle.com/mathurinache/world-happiness-report for data on happiness parameters. The data set on suicides included yearly data from 1985 till 2016 on fields: (i) Country; (ii) Year; (iii) Gender; (iv) Age group (5-14, 15-24, 25-34 35-54, 55-74 and over 75 years); (v) No. of suicides; (vi) Country’s population; (vii) HDI (Human Development Index); (viii) GDP; and (ix) Generation (Generation Z, Millennials, generation X and Silent). Data for many countries did not extend to the entire range i.e. 1985-2016. The first part of the study focused on using Machine Learning algorithms for identifying predictors for suicide. Predictors here are those factors (features) that play an important role in determining suicide rate in the country. Next, data from world happiness report was combined with suicide data to develop an understanding of country specific factors that can help address the challenge of suicides. Once again, machine learning was applied for the purpose using IBM SPSS Modeler. World happiness report provided data on (i) Country; (ii) Region (for country); (iii) Happiness Rank; (iv) Happiness Score (scaled: 2.9-7.54); (v) Economy (scaled: 0.09-1.74); (vi) Family (scaled: 0.43-1.61); (vii) Health (scaled: 0.01-0.95); (viii) Freedom (scaled: 0.03-0.66); (ix) Generosity (scaled: 0-0.84); (x) Govrnment_trust (scaled: 0-0.46); and (xi) Dystopia_residual (scaled: 0.38-2.9) with dystopia depicting the very opposite of utopia.

### ANALYSIS OF DATA AND SUMMARY OF FINDINGS

Data for the study was taken from the kaggle site: https://www.kaggle.com/russellyates88/suicide-rates-overview-1985-to-2016. The data set comprised 27820 records on about 6.75 Million suicides spread across 101 countries in the period from 1987 to 2016. The fields in the dataset included: Country, Year, Gender, Age (six age groups of 5-14, 15-24, 25-34, 35-54, 55-74 and 74+ years), No. of suicides, Population, HDI index, GDP and Generation (i.e. Boomers, Generation X, Silent Generation, or G.I.Generation). The data was analysed using IBM SPSS Modeler. Multiple ML models were run so as to put together the findings/result of all models. The models run included: (i) Neural network; (ii) Regression; (iii) Random Forest; (iv) XG Boost Tree; (v) XG Boost-AS; (vi) CHAID; (vii) GLMM; (viii) Generalized Linear; (ix) Random Trees; (x) Tree-AS; and (xi) Auto Numeric (ensemble) Model. These findings are presented below:

**Fig. 8:**
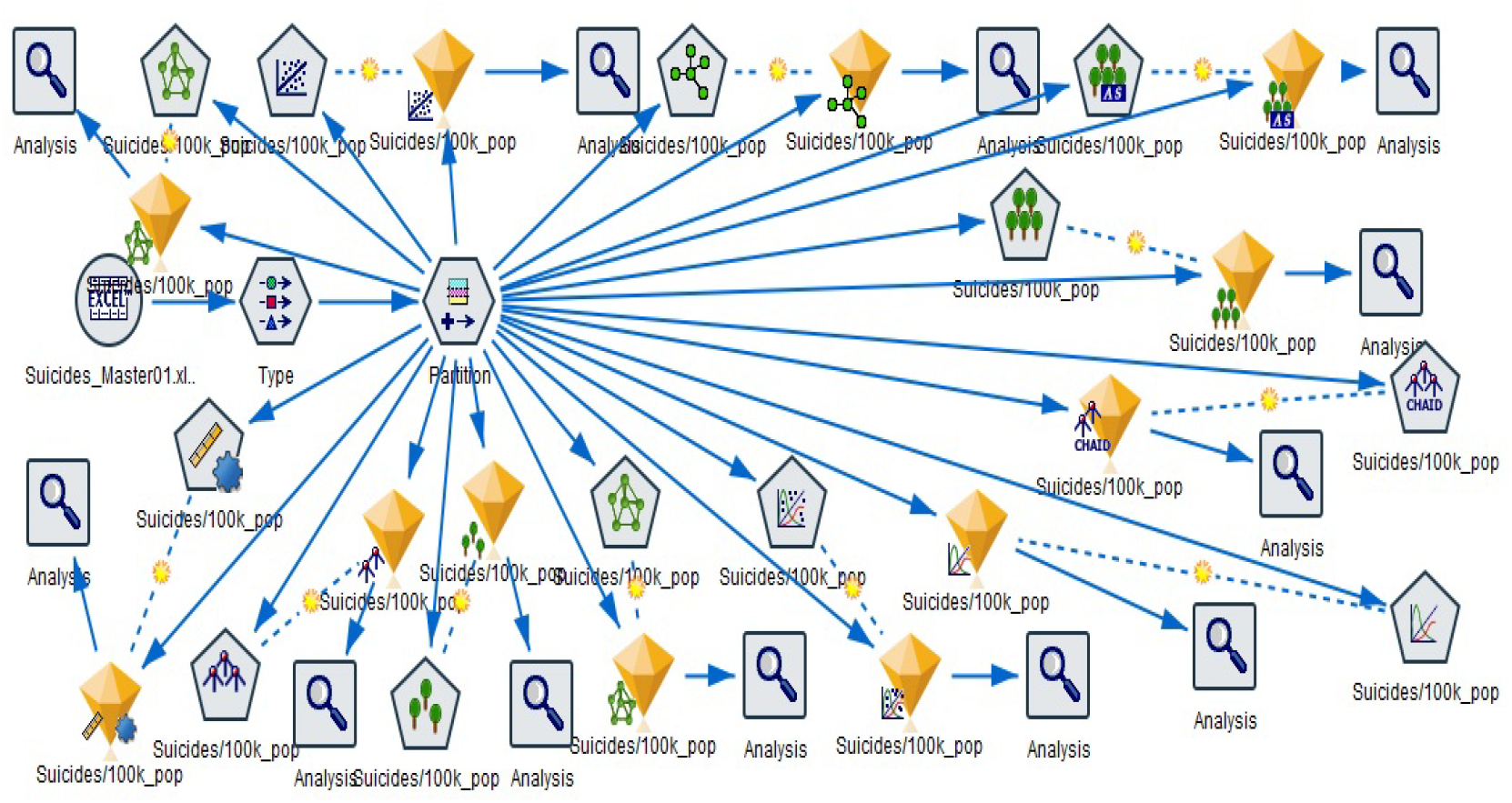
Pictorial Summary: ML Models (Suicide across Countries)

All ML models were deployed with the same target variable, namely, ‘Suicides/100000 Population’ and used the same ratio of train: test split i.e 75:25. Findings from these machine learning models are presented next: Neural Network: The hidden layer used had 13 neurons besides the bias unit. This Neural Network model gave an accuracy of 76.3% through a Multi-Layer perceptron as shown below.

**Fig. 9:**
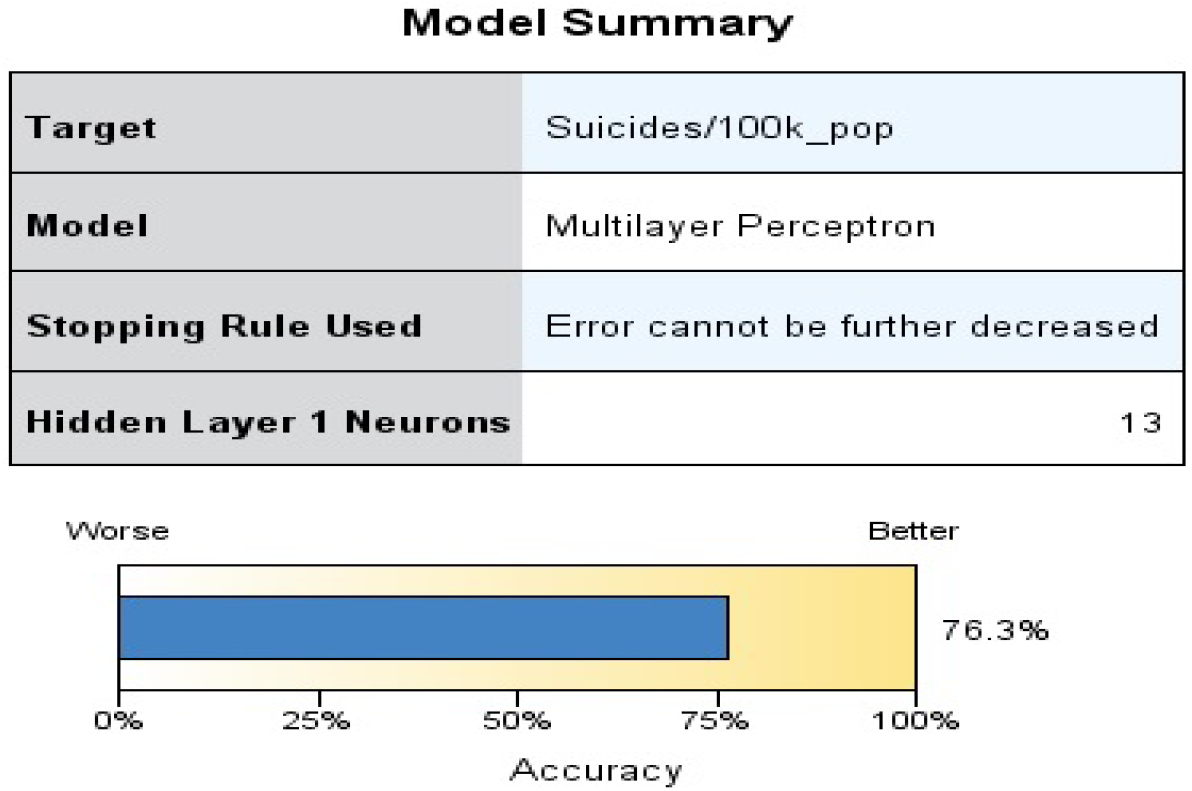
Accuracy of Neural Network Model (One Hidden Layer)

A schematic representation of the network with one hidden layer carrying 13 nodes (besides the bias unit) is presented below.

**Fig. 10:**
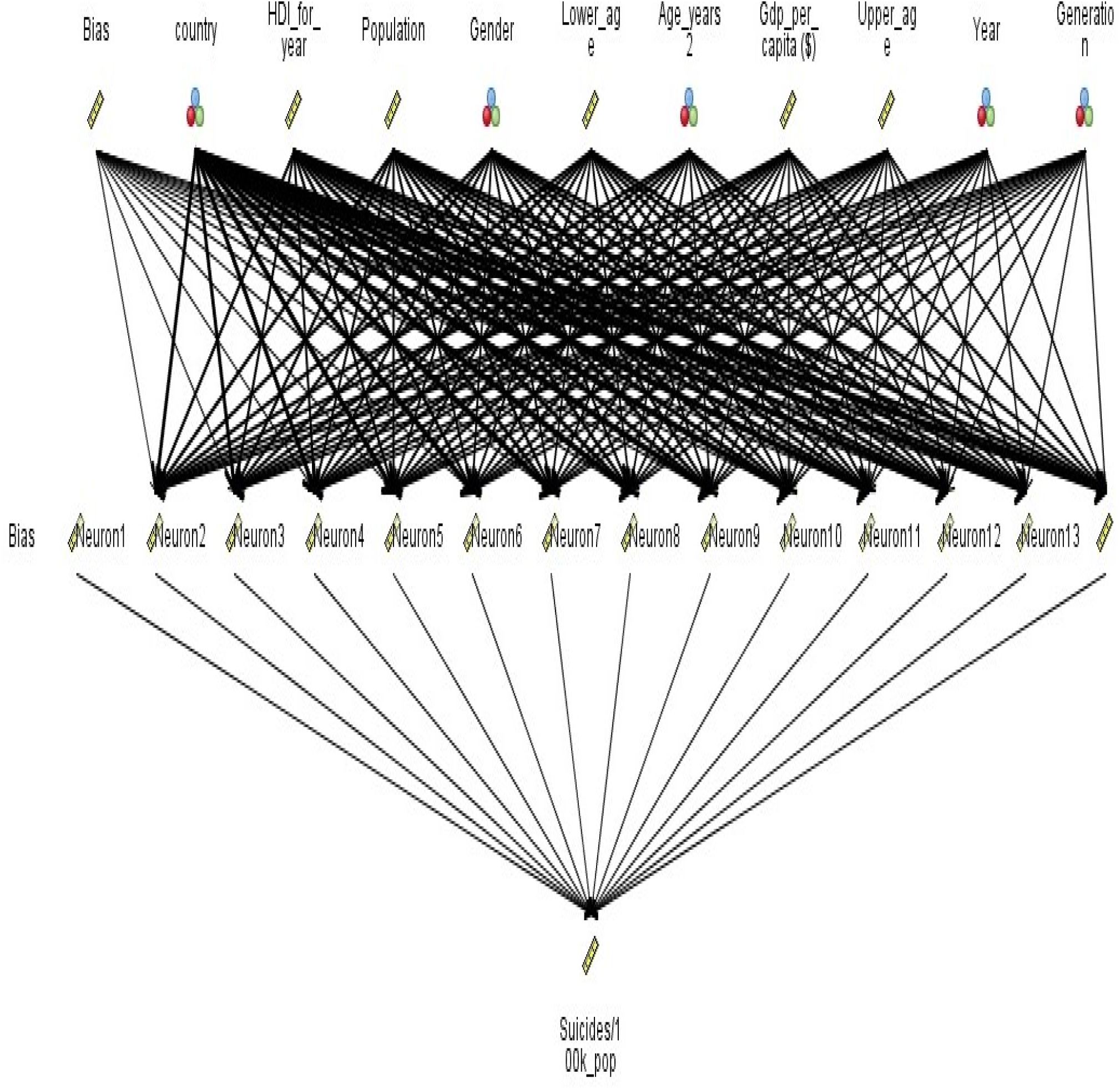
Fully Connected Neural Network with one hidden layer.

Predictor importance of different models was seen to differ and is presented below:

**Fig. 11 (a):**
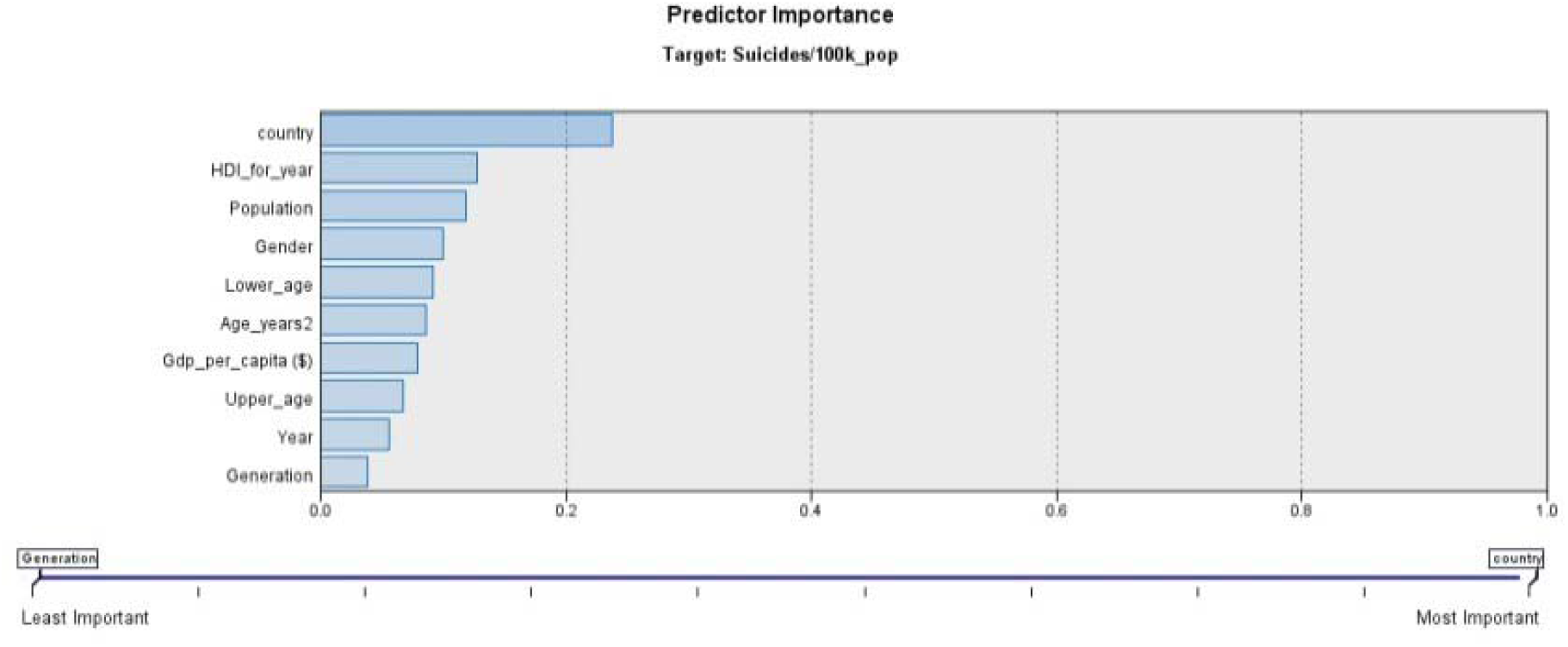
Predictor Importance: Neural Network (One Hidden Layer)

The main predictors are (i) Country: 24%; (ii) HDI: 13%; (iii) Population: 12% and (iv) Gender: 10%.

**Fig. 11 (b):**
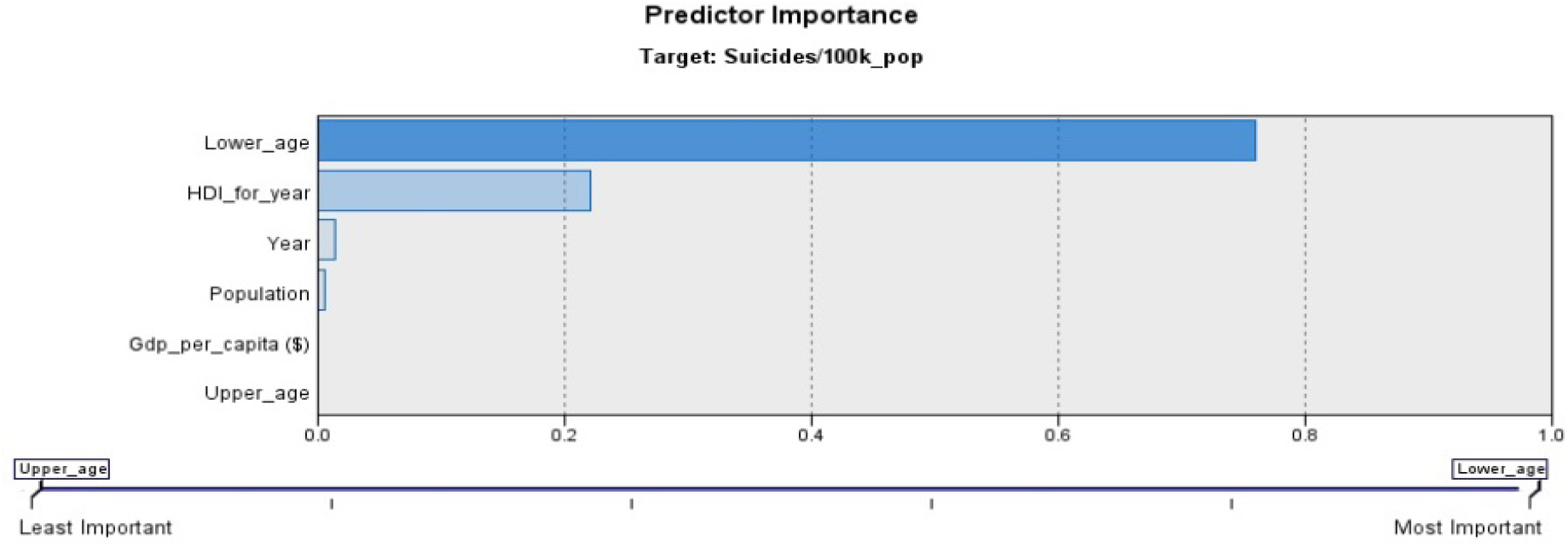
Predictor Importance: Regression Model.

The main predictors are (i) Lower_age (from 6 categories of age groups): 76%; and (ii) Human Development Index or HDI: 22%.

**Fig. 11 (c):**
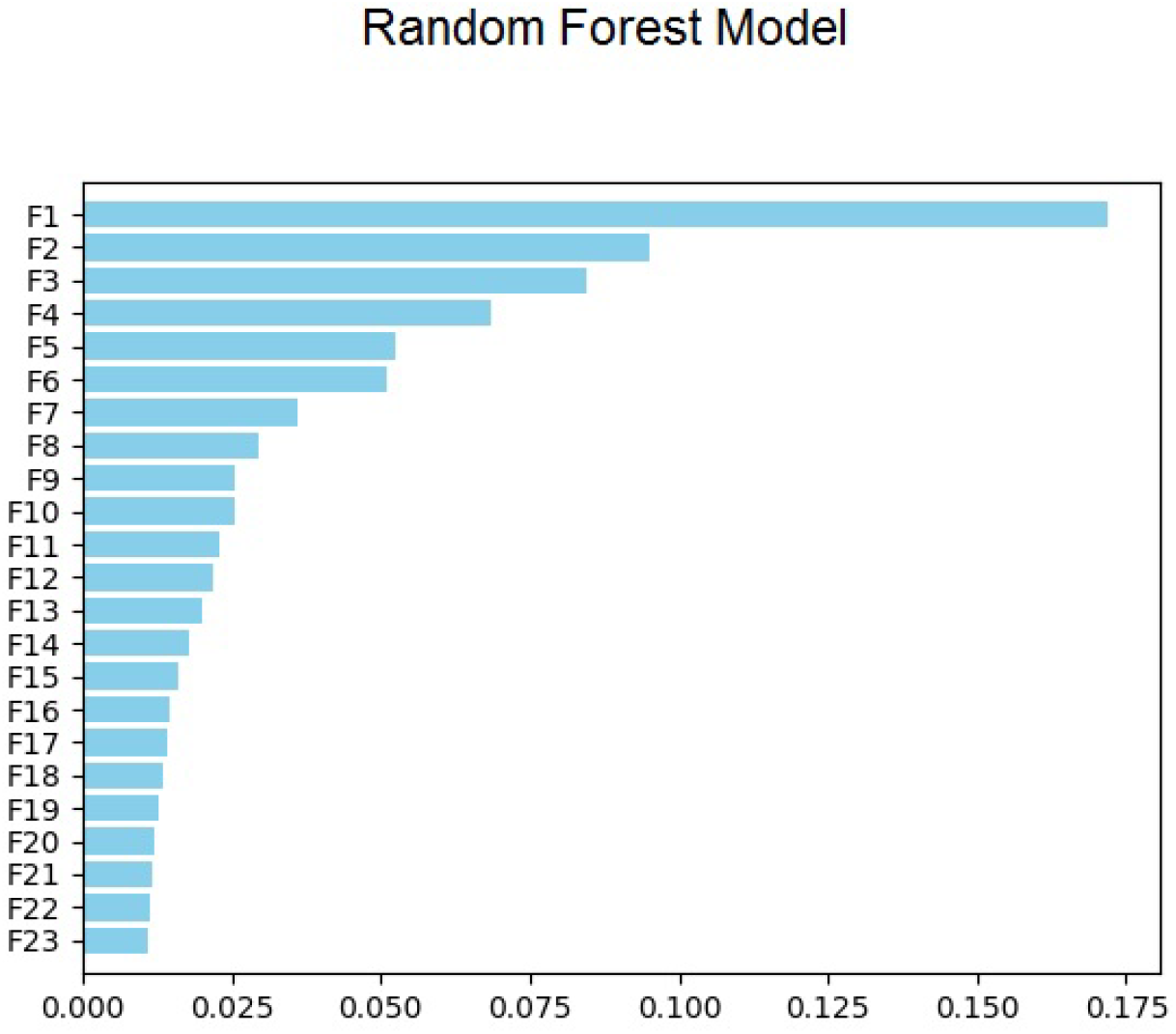
Predictor Importance: Random Forest Model.

The key predictors are: (i) Gender: Over 17%; and (ii) Lower_age (from 6 categories of age groups): About 10%.

**Fig. 11 (d):**
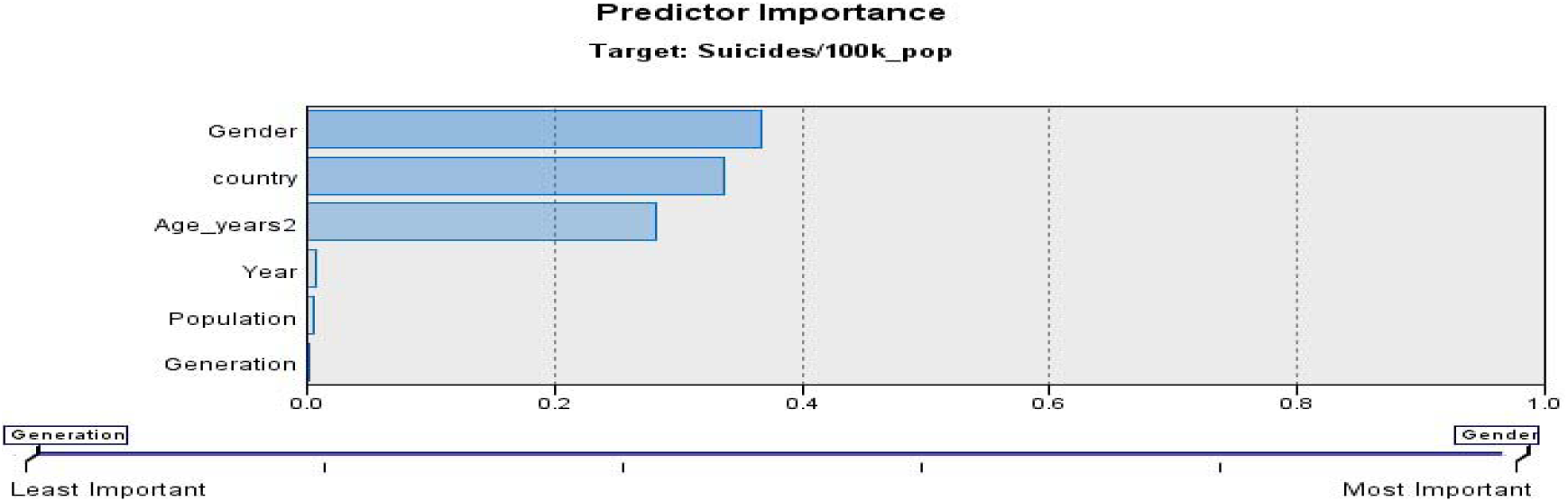
Predictor Importance: CHAID Model.

The key predictors are: (i) Gender: 37%; (ii) Country: 34%; and (iii) Age (group): 28%. The information given above on predictors is summarized as below:

Assigning a weightage of 3 to the most important predictor, 2 to the second in importance predictor and 1 to the third in Table 1 leads to Gender and Age emerging as the key predictors. Variation of suicide rate with respect to age shows a higher rate in the older age groups across the world as shown in the graphs below. For better presentability, the number of countries was limited in each graph and countries presented in decreasing order of suicide rate. Countries were categorized into four groups based on suicide rates, each category approximating a quartile.

**Table 1:** Identifying the Key Predictors (factors) of Suicide based on Suicide data.

| Predictor | Most Important | 2<sup>nd</sup> in importance | 3<sup>rd</sup> in importance |
| --- | --- | --- | --- |
| Gender | CHAID<br>Random Forest |  |  |
| Country | Neural Net (1 Hidden layer) | CHAID |  |
| HDI |  | Neural Net (1 hidden layer)<br>Regression |  |
| Population |  |  | Neural Net (1 hidden layer) |
| Age* | Regression | Random Forest | CHAID |

\* Age here presents a combined view of age group, lower-end and upper-end of age group.

**Fig. 12 (a).**
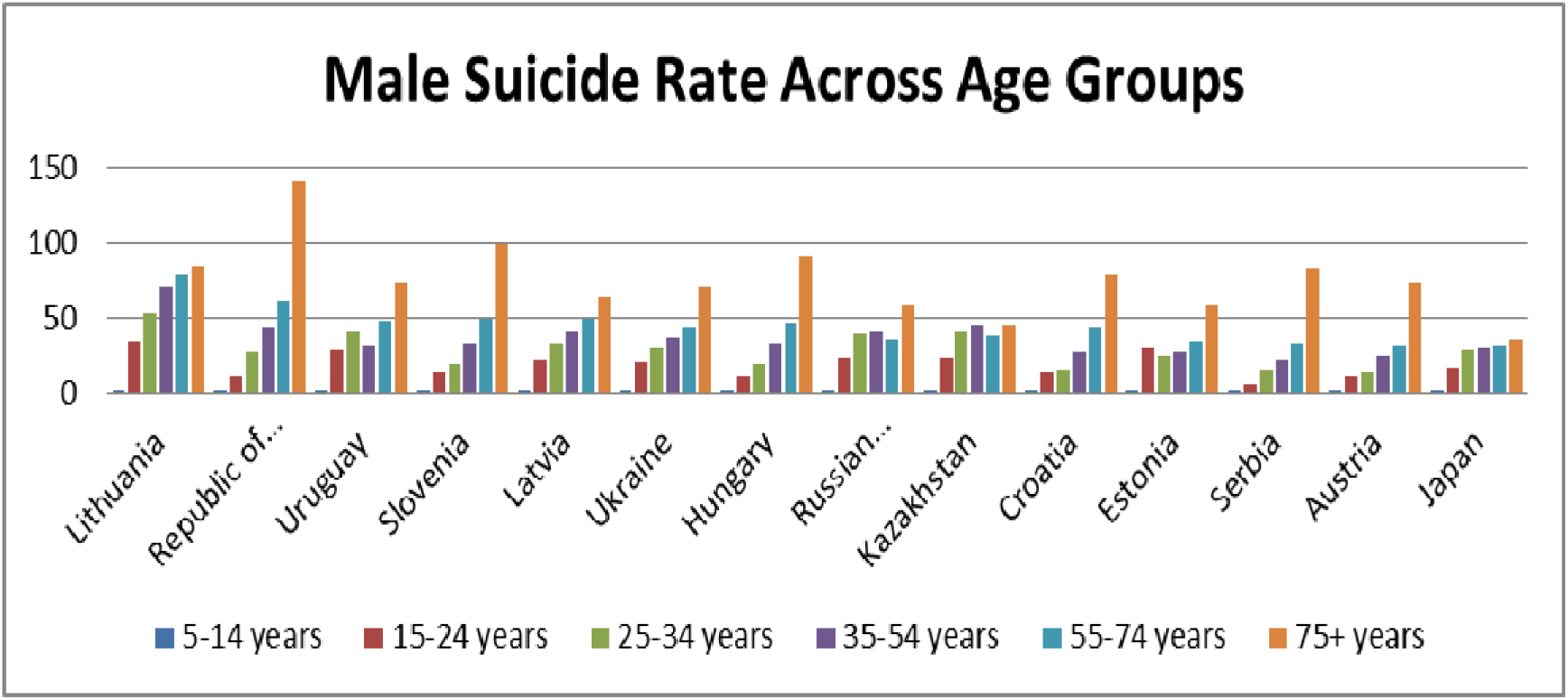
Suicide Rate across Age Groups (Male) in Countries (14 highest suicide rates)

The graph above leaves no doubts about the higher seriousness of the problem in older generations.

**Fig. 12 (b).**
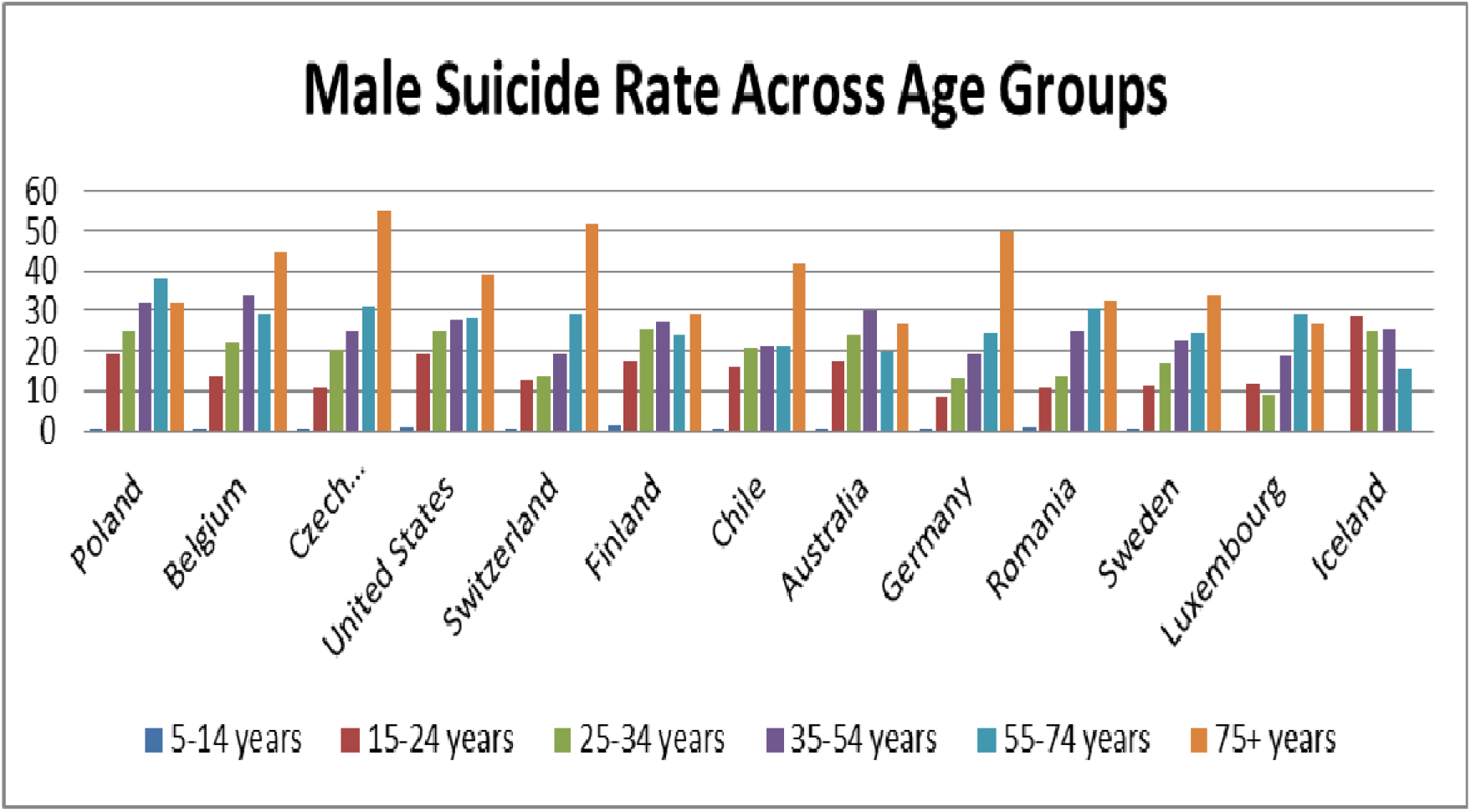
Suicide Rate across Age Groups (Male) in Countries (14 high suicide rates)

Once again, the bars are longest for the higher age group males on almost all countries above with relatively less populated countries like Luxembourg and Iceland showing a slightly different pattern.

**Fig. 12 (c).**
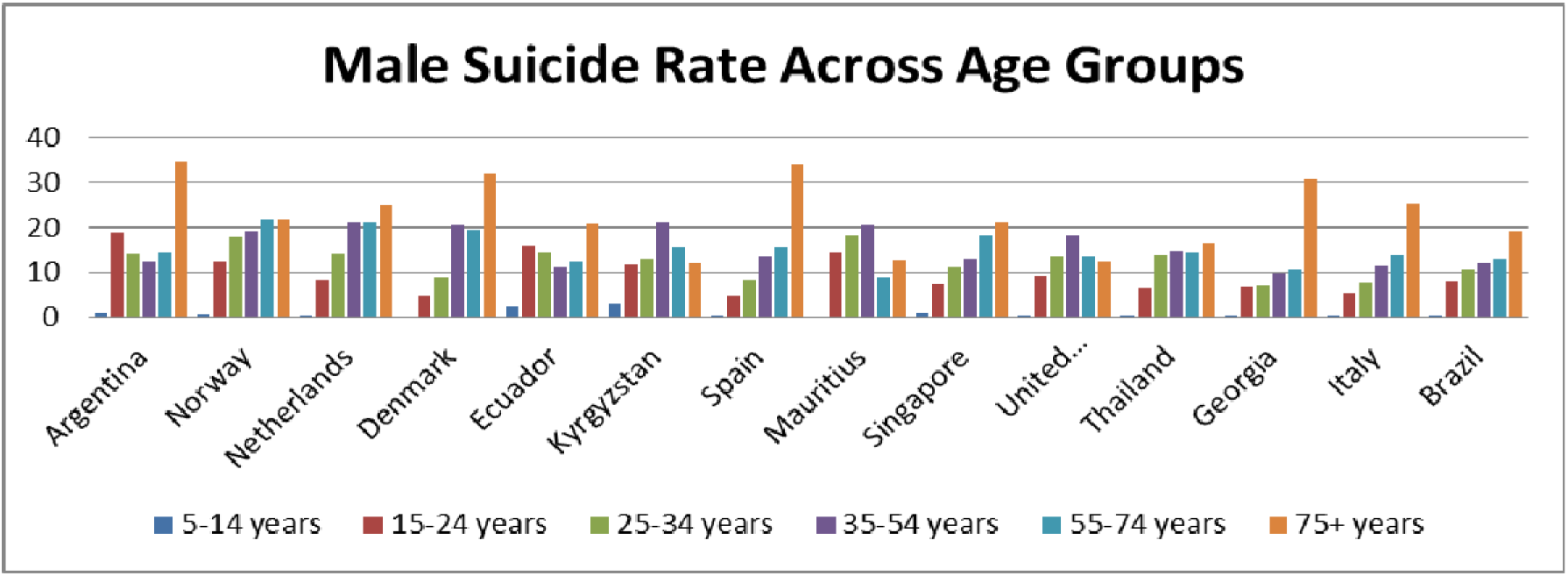
Suicide Rate across Age Groups (Male) in Countries.

Suicide rate here demonstrates the same pattern with higher rate in the age groups 55-74 and 75+.

**Fig. 12 (d).**
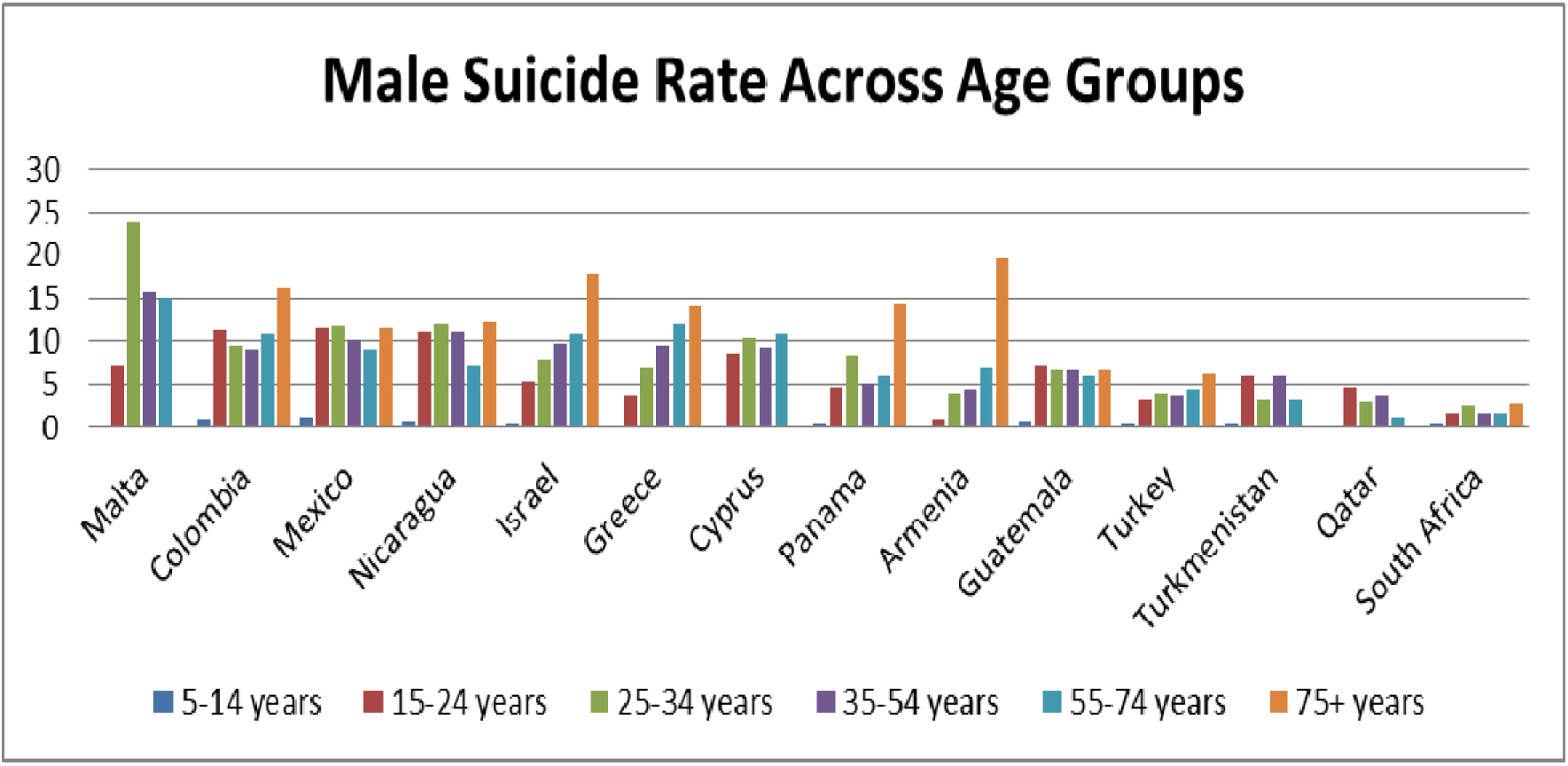
Suicide Rate across Age Groups (Male) in Countries (Lowest suicide rate)

In countries with the lowest suicide rates, one can observe a different pattern that is not necessarily tilted towards higher rate among the higher age groups.

Next, a review of suicides among female members of populations also reveals a similar trend as for males with minor differences in countries that fall in the lower 50% as far as suicide rate goes.

**Fig. 13 (a).**
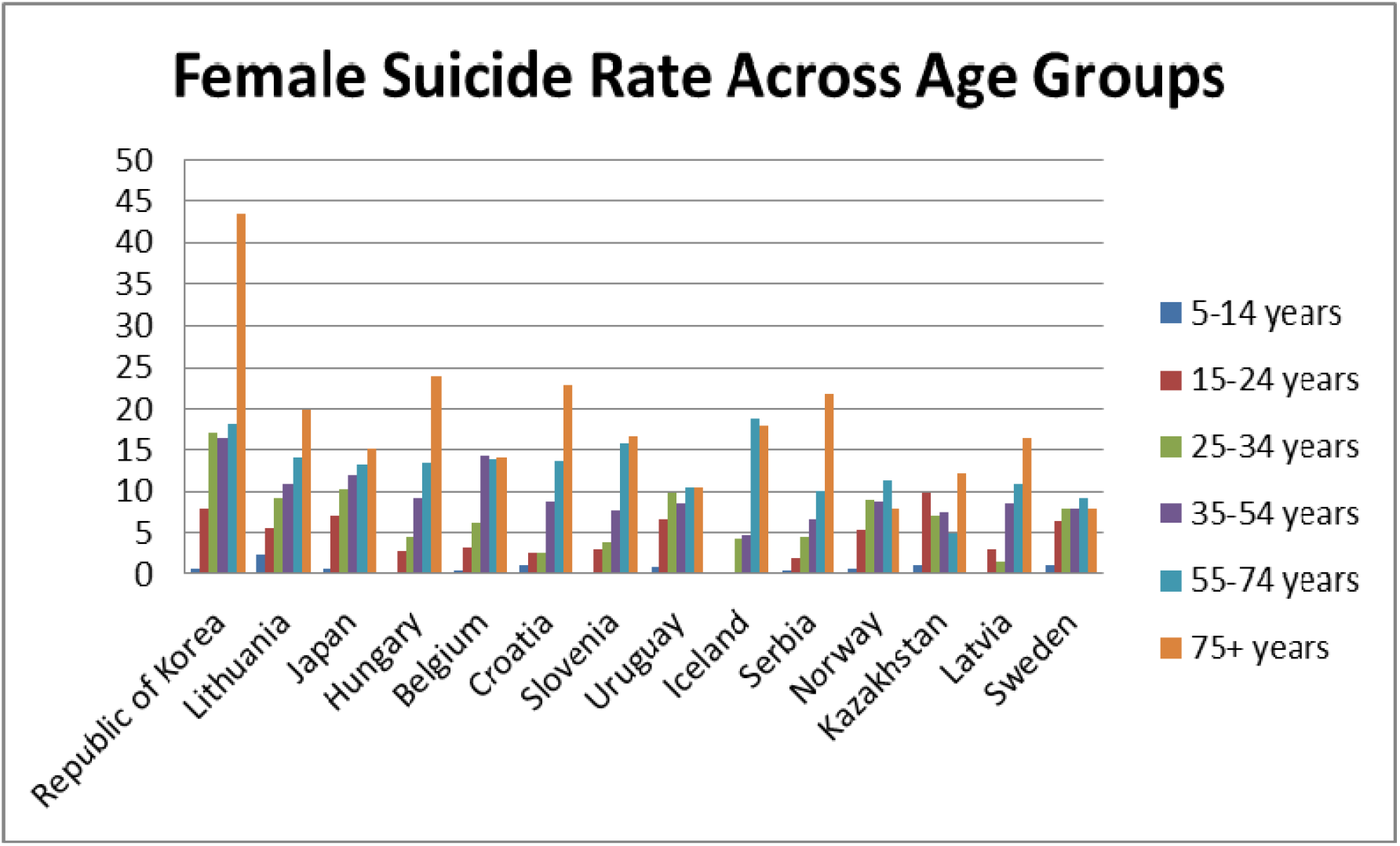
Suicide Rate across Age Groups (Female) in Countries (14 Highest suicide rates)

The graphical representation above highlights the seriousness of the problem of suicides among the old.

**Fig. 13 (b).**
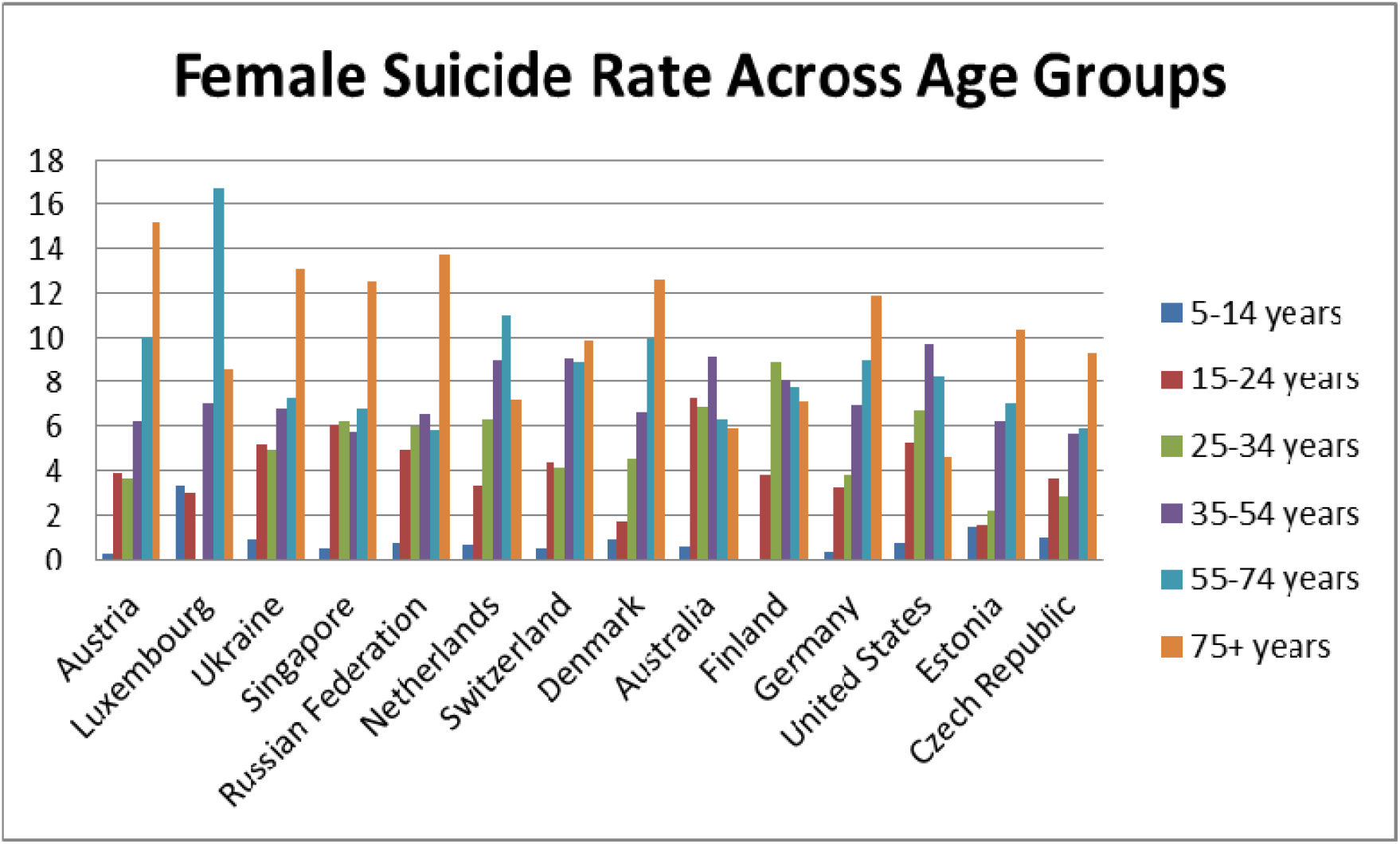
Suicide Rate across Age Groups (Female) in Countries (14 high suicide rates)

Barring the relatively thinly populated Luxembourg, in all cases the 75+ years category is most seriously affected by suicides. Luxembourg showed a slight variation in pattern even among the male members of population suicides with the 55-74 age group experiencing higher suicide rates than the 75+ age group.

**Fig. 13 (c).**
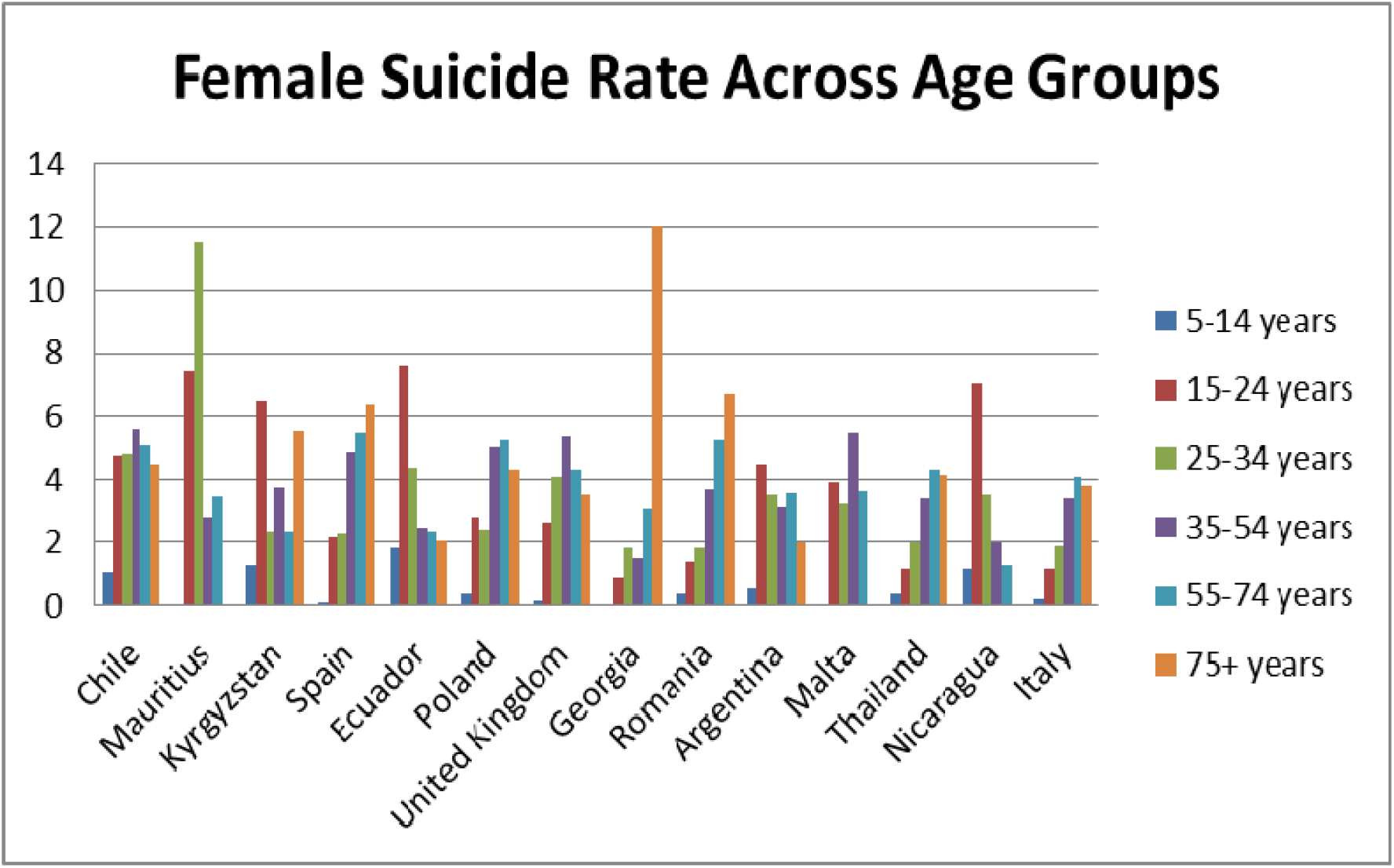
Suicide Rate across Age Groups (Female) in Countries.

One can see a changed pattern here with several countries facing higher suicide rates in lower age groups.

**Fig. 13 (d).**
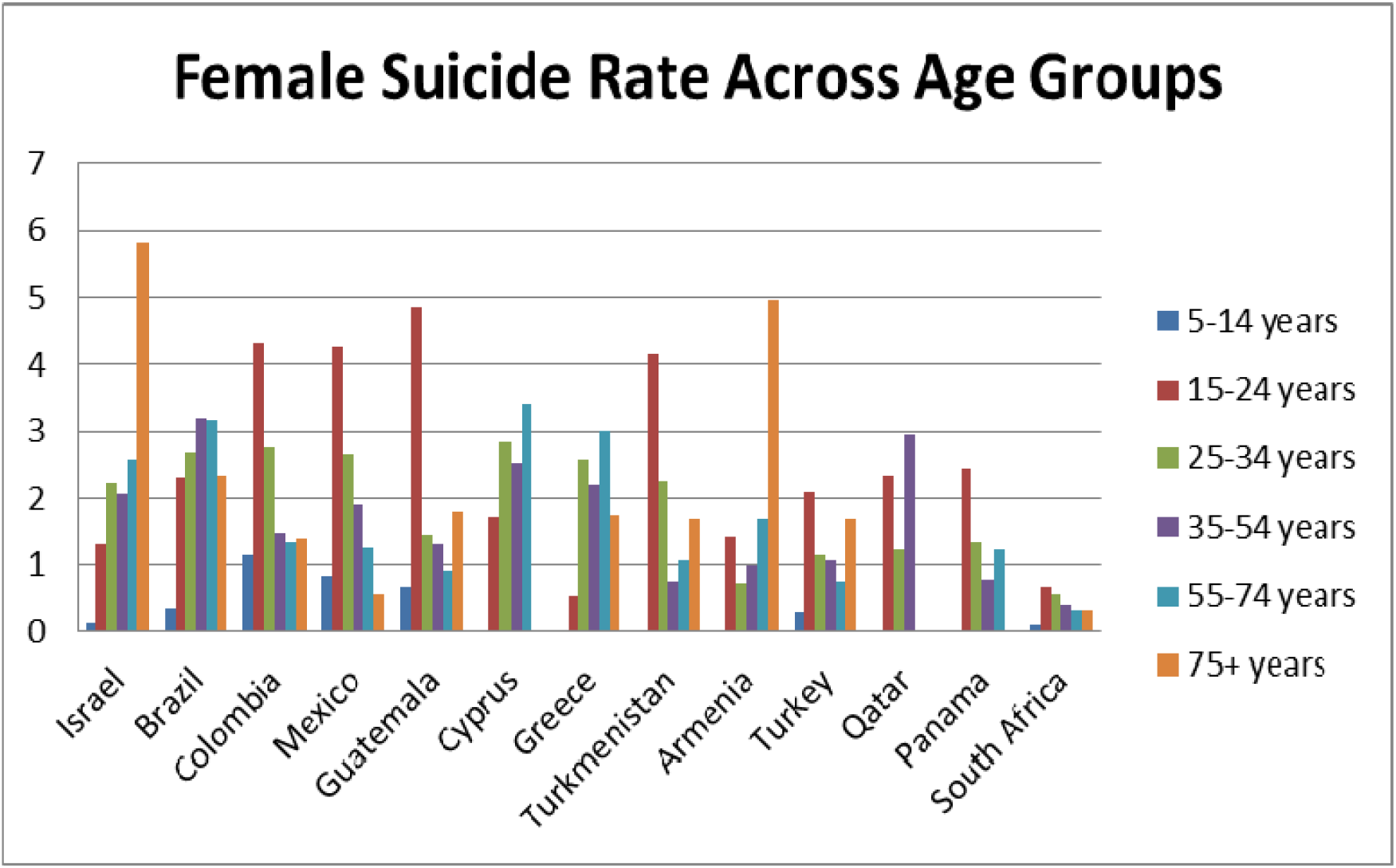
Suicide Rate across Age Groups (Female) in Countries (Lowest suicide rate)

It can be seen that while higher age groups are certainly more at risk of suicides for both genders, the pattern is particularly more pronounced when suicide rate in the country is on the higher side. In countries with lower suicide rates, the spread is more among female members of the population as compared to males. Variation in suicide rate based on gender is presented below for countries with the highest suicide rates.

**Fig. 13 (e).**
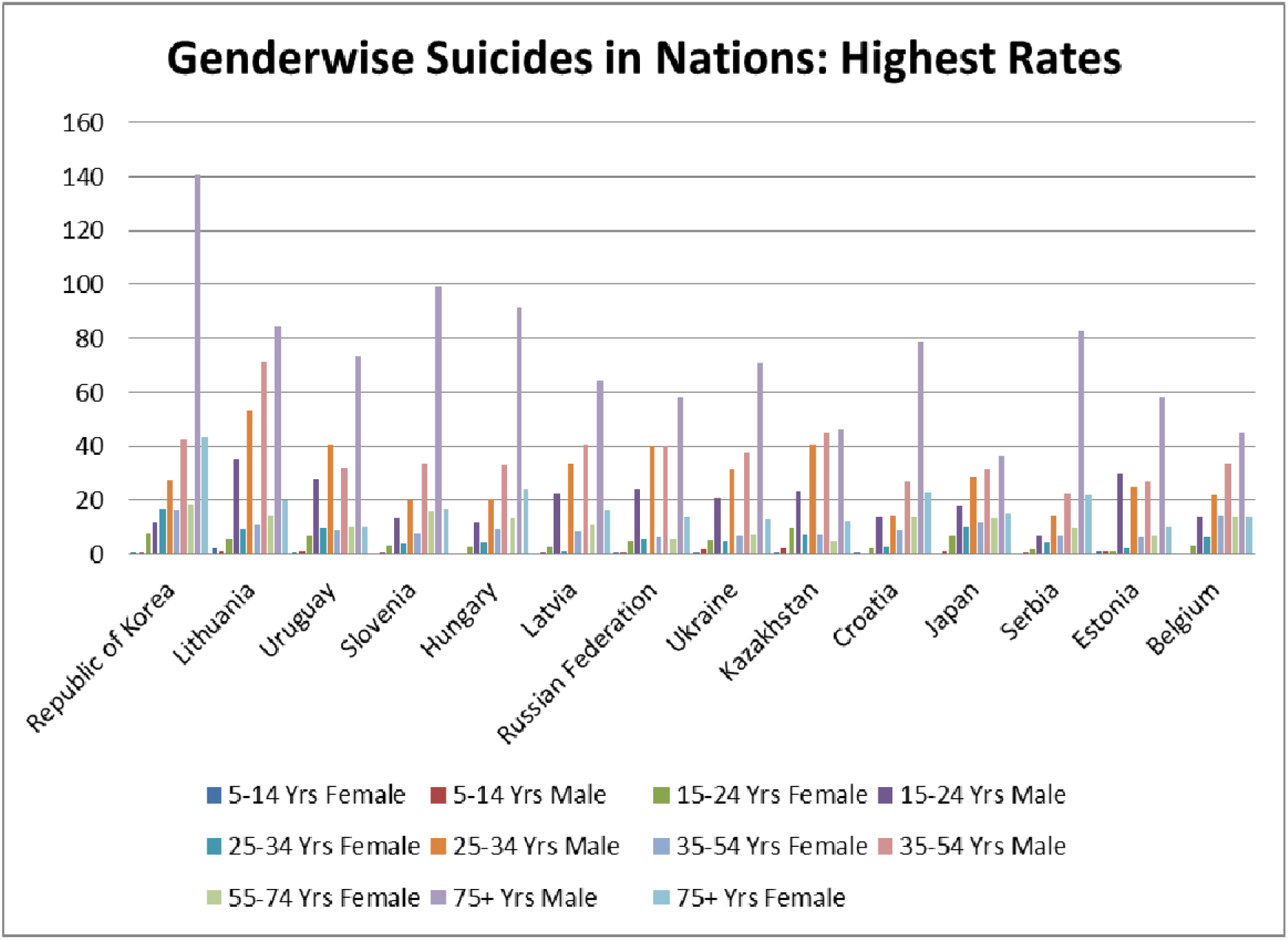
Suicide Rate across Age Groups (Male and Female) in Countries (Highest suicide rate)

The graphical representation above brings out the higher suicide rates among higher age groups and also among males, particularly in the higher age groups. Combining these suggests that while Gender, Age and Country are important predictors for suicides, Gender and Age are unidimensional characteristics i.e. they represent one specific feature that takes a value that is indisputable, ‘Country’, on the other hand, represents the host of features that impact all eg. prevailing social values, practices, pressures besides other country specific features. These form the subject of subsequent discussions below with focus on countries that reported a population of over a million (in the dataset) in latest year (2014 or 2015). Once again, for presentability, the number of countries was limited in each graph.

**Fig. 14 (a):**
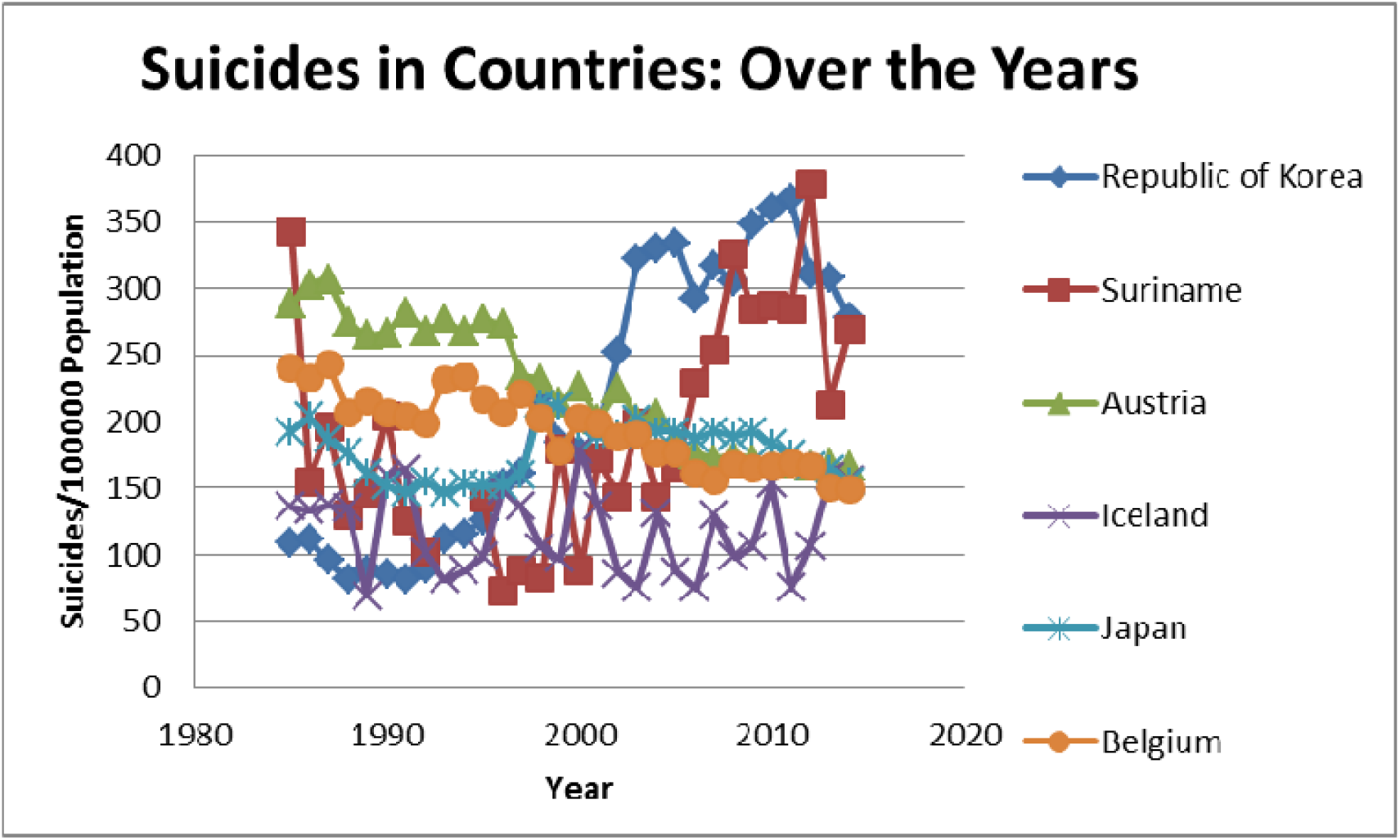
Suicides in individual countries over the years.

Austria is among the countries that have shown a downward trend as regards suicides with Belgium also showing improvements. Other countries have not seen similar improvements.

**Fig. 14 (b):**
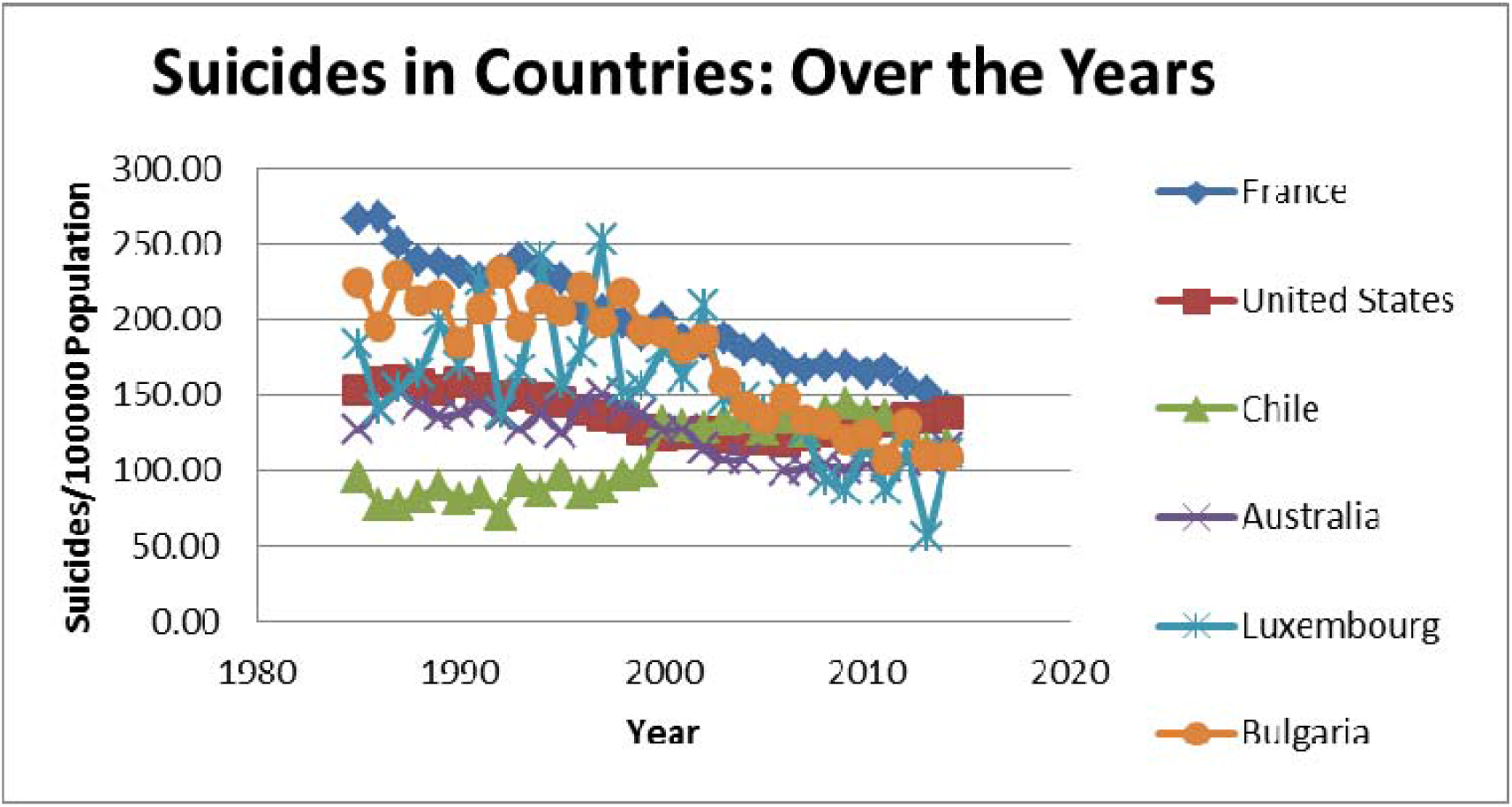
Suicides in individual countries over the years.

In the graph above, France, Bulgaria and Luxembourg show a downward trend while Chile has seen a deterioration and USA showing relative stability in trend.

**Fig. 14 (c):**
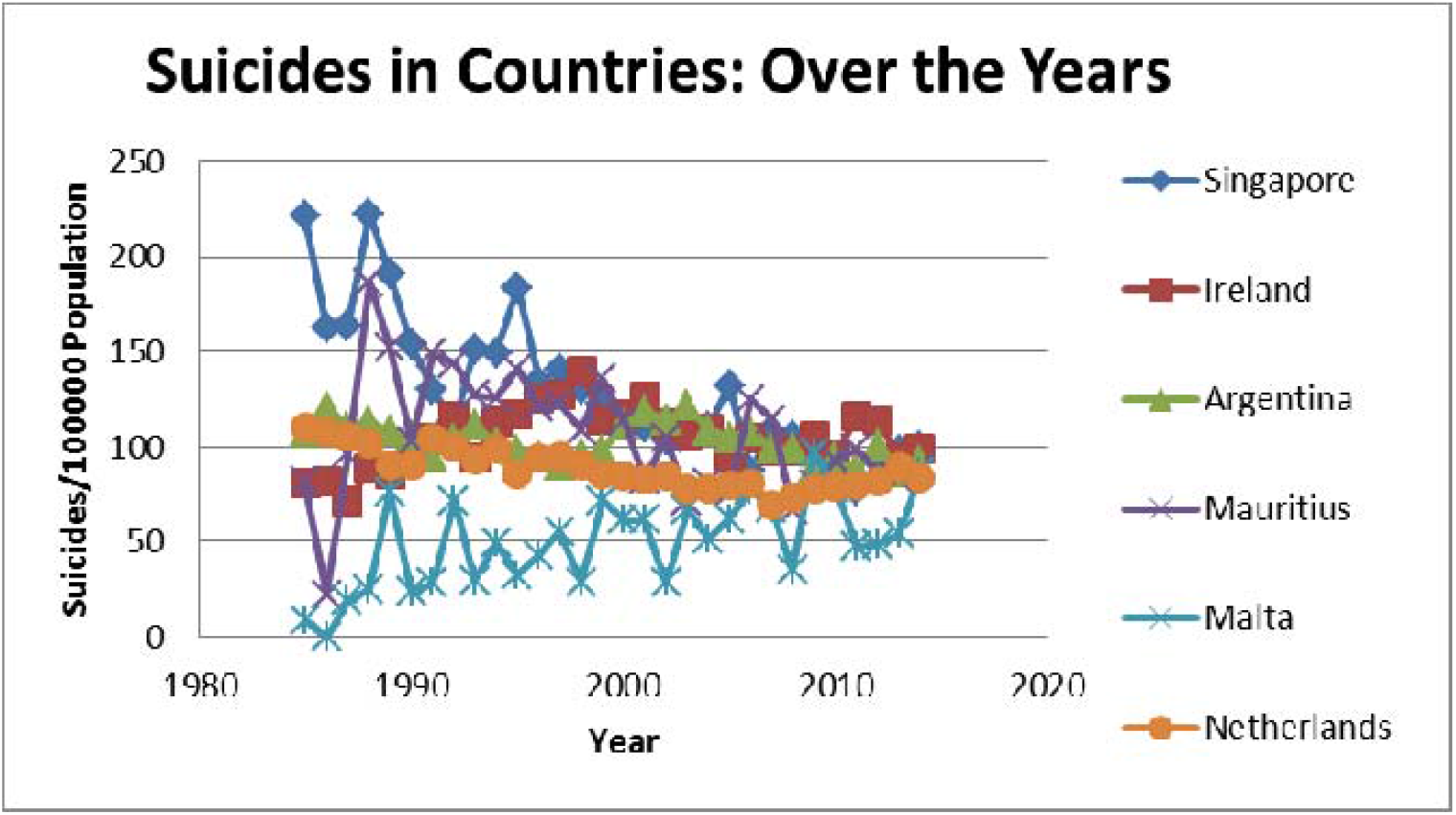
Suicides in individual countries over the years.

The graph above shows shows a downward trend in Singapore, relative stability in the trends in Ireland, Argentina, Mauritius and Netherlands while Malta, with a population much less than a million in 2015, shows a deteriorating trend.

**Fig. 14 (d):**
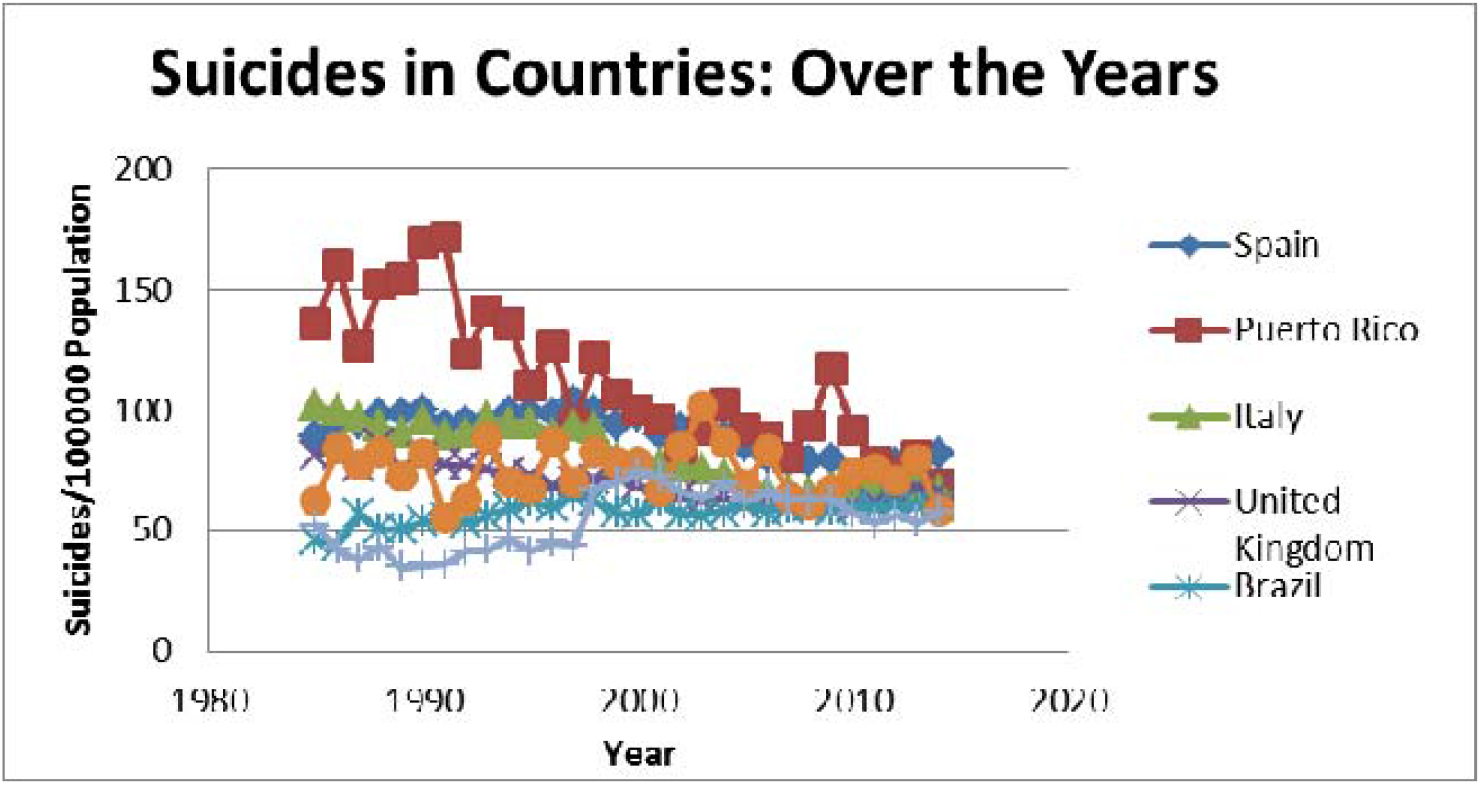
Suicides in individual countries over the years.

While Peurto Rico shows a clear downward trend as regards suicides, Spain, Italy, UK and Costa Rica show relative stability in suicides and Brazil as well as Columbia have seen rising incidence of suicides.

**Fig. 14 (e):**
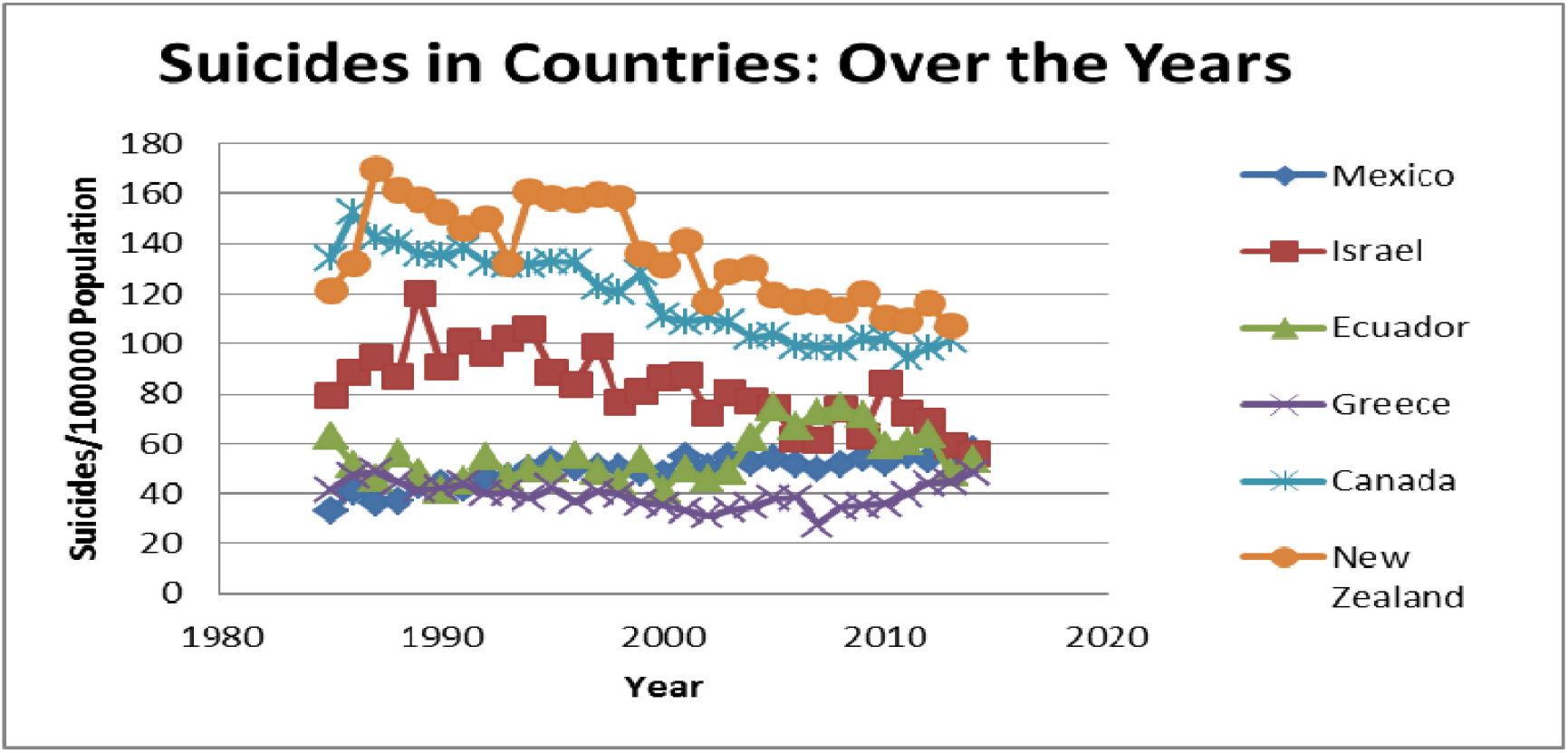
Suicides in individual countries over the years.

The graph above shows increasing rate in suicides in Mexico but other countries, namely, Israel, Equador, Greece and New Zealand show relative stability and Canada shows improving trends. To summarize, countries with a population above 1 Million and deteriorating trends in suicides include Brazil, Chile, Columbia, Mexico and Republic of Korea while countries that have seen improvements in suicide rates include Austria, Belgium, Bulgaria, Canada, France, Puerto Rico and Singapore. Israel and New Zealand have also witnessed some improvements. The fact that various Machine Learning Models do not include Per Capita GDP as among the top three predictors shows its relatively smaller role. This is further established by the upward trend over years in GDP for these countries irrespective of increasing or falling suicide rates as shown in the figure below.

**Fig. 15 (a):**
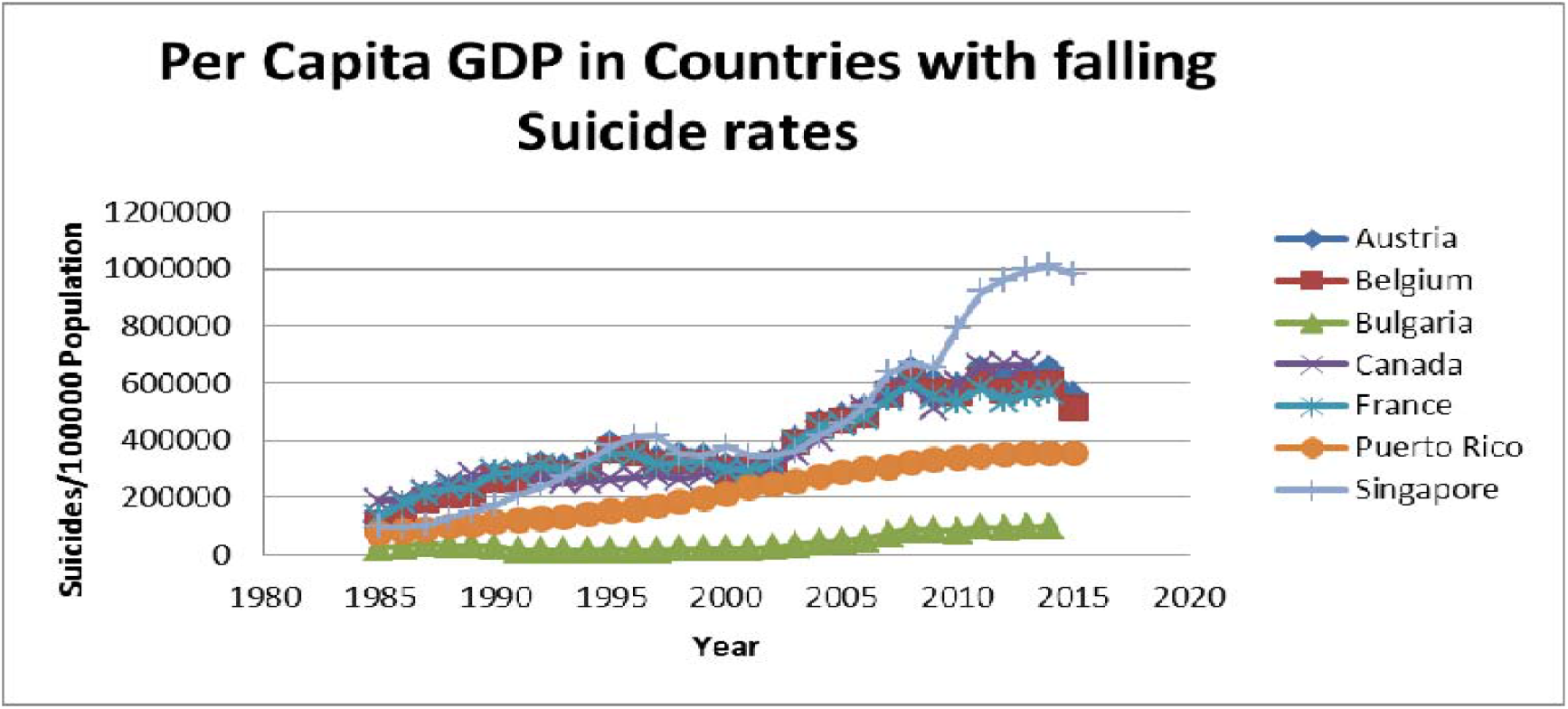
Per Capita in Countries with falling suicide rates.

**Fig. 15 (b):**
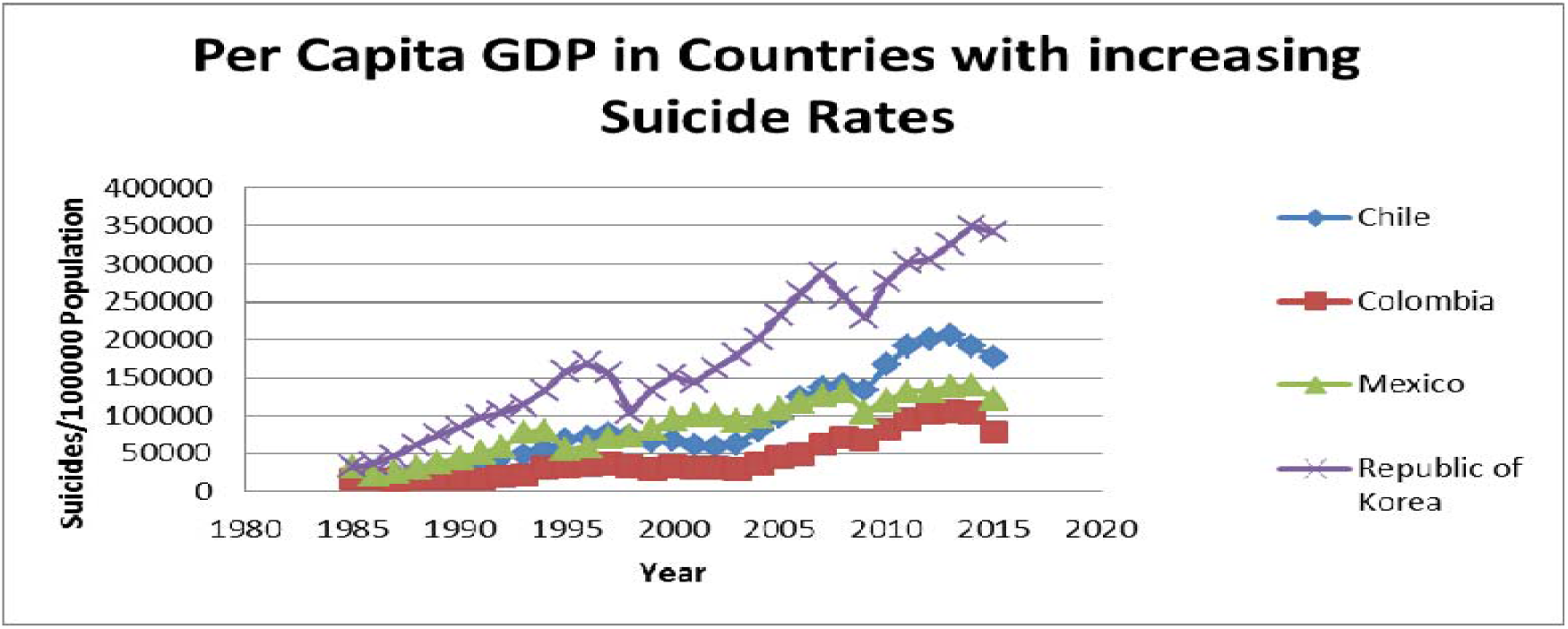
Per Capita in Countries with falling suicide rates.

To understand the linkage between country specific factors and suicides, data from the world happiness report was merged with suicide data and various machine learning models applied to get results enumerated below. As the common period for suicides data and happiness report was only the year 2015, further analysis relates to merging suicide data with the happiness report data for 2015. This merging provided 660 data points for 55 countries (12 per country with one data point for each gender all six age categories, namely, 5-14 years, 15-24 years, 25-34 years, 35-54 years, 55-74 years and 75+ years). The following machine learning models were applied to the merged data set: (a) Linear; (ii) XG-Boost Tree-AS; (iii) CHAID; (iv) Generalized Linear; (v) Deep Neural Network with two hidden layers; (vi) Random Forest; (vii) Deep Neural Network with one hidden layer; (viii) Regression; (ix) XG-Boost Tree; (x) Random Trees; and (xi) Auto numeric (ensemble) model. Summary

**Fig. 16:**
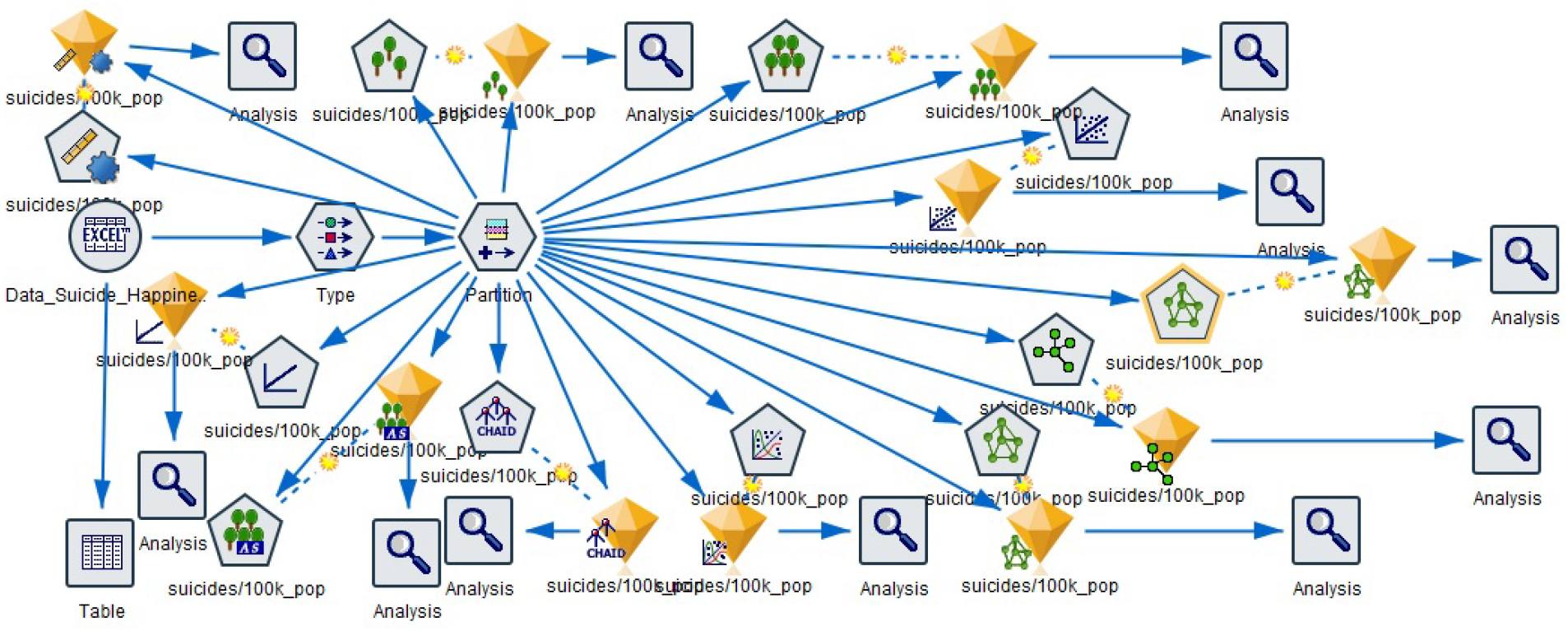
Pictorial Summary of ML Models Run on Merged Happiness and Suicide Data.

These models provided the following as key predictors of suicide:

**Fig. 17 (a):**
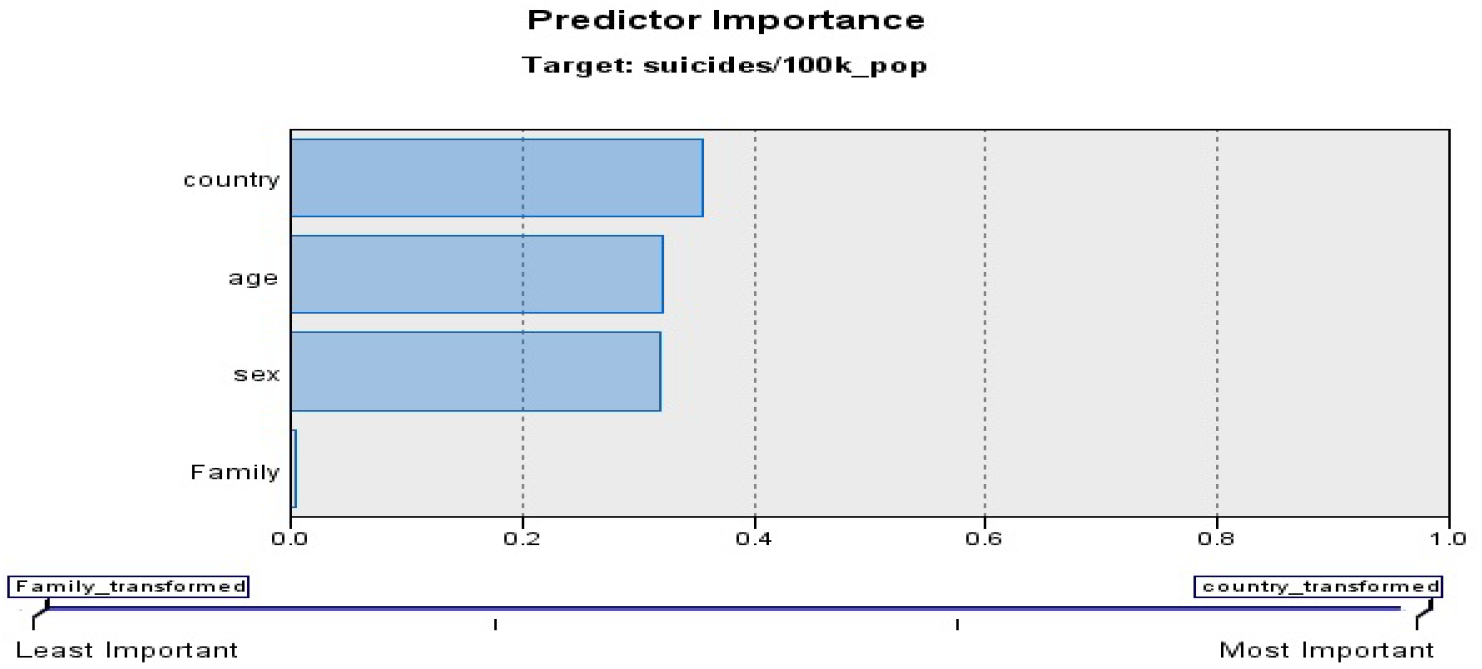
Predictor Importance: Linear Model.

The key predictors are: (a) Country: 36%; (b) Age (group): 32%; and (c) Gender: 32%

**Fig. 17 (b):**
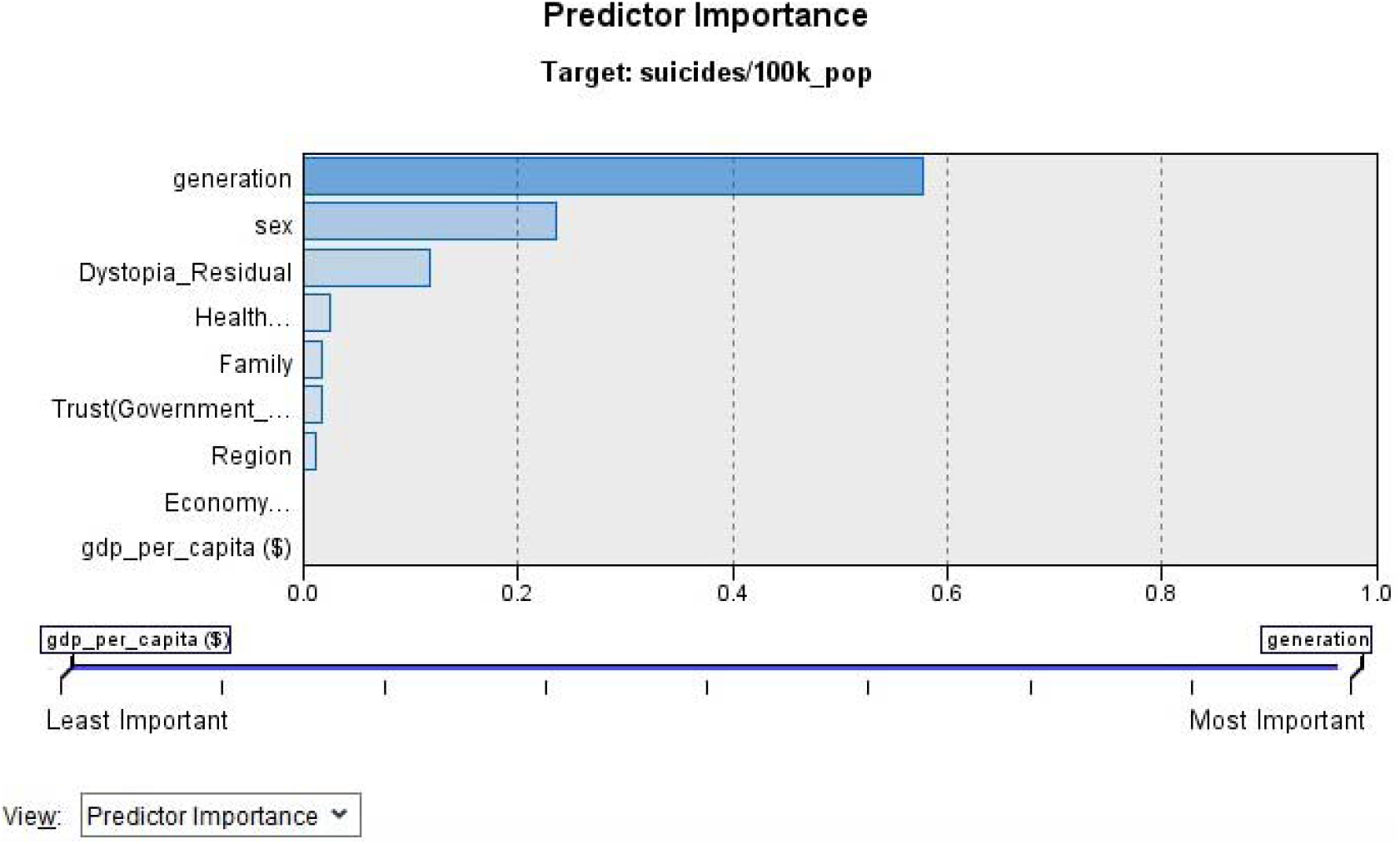
Predictor Importance: CHAID Model.

The main predictors are: (a) Generation: 58%; (b) Gender: 24%; and (c) Dystopia Residual: 12%.

**Fig. 17 (c):**
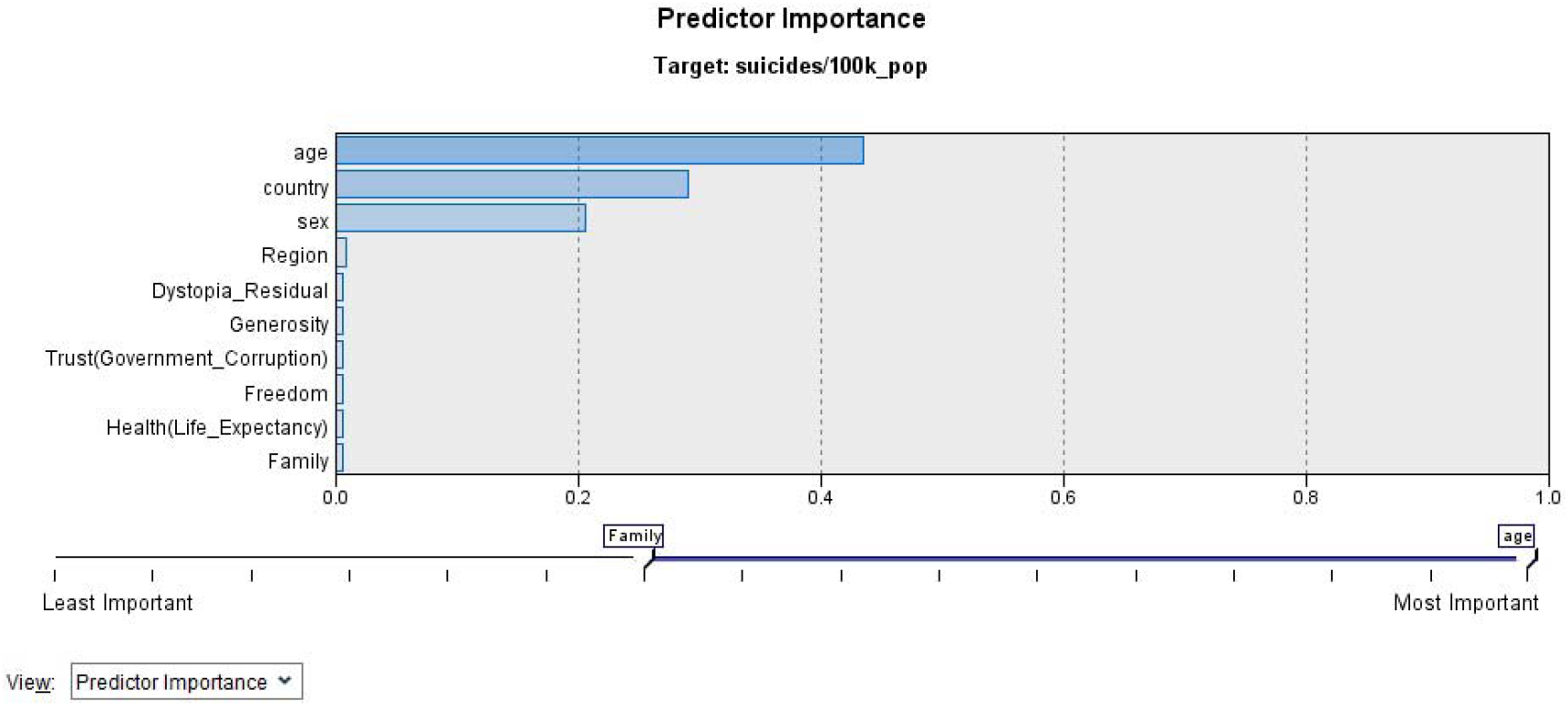
Predictor Importance: Generalized Linear Model.

The key predictors are: (a) Age (group): 44%; (b) Country: 29%; and (c) Gender: 20%

**Fig. 17 (d):**
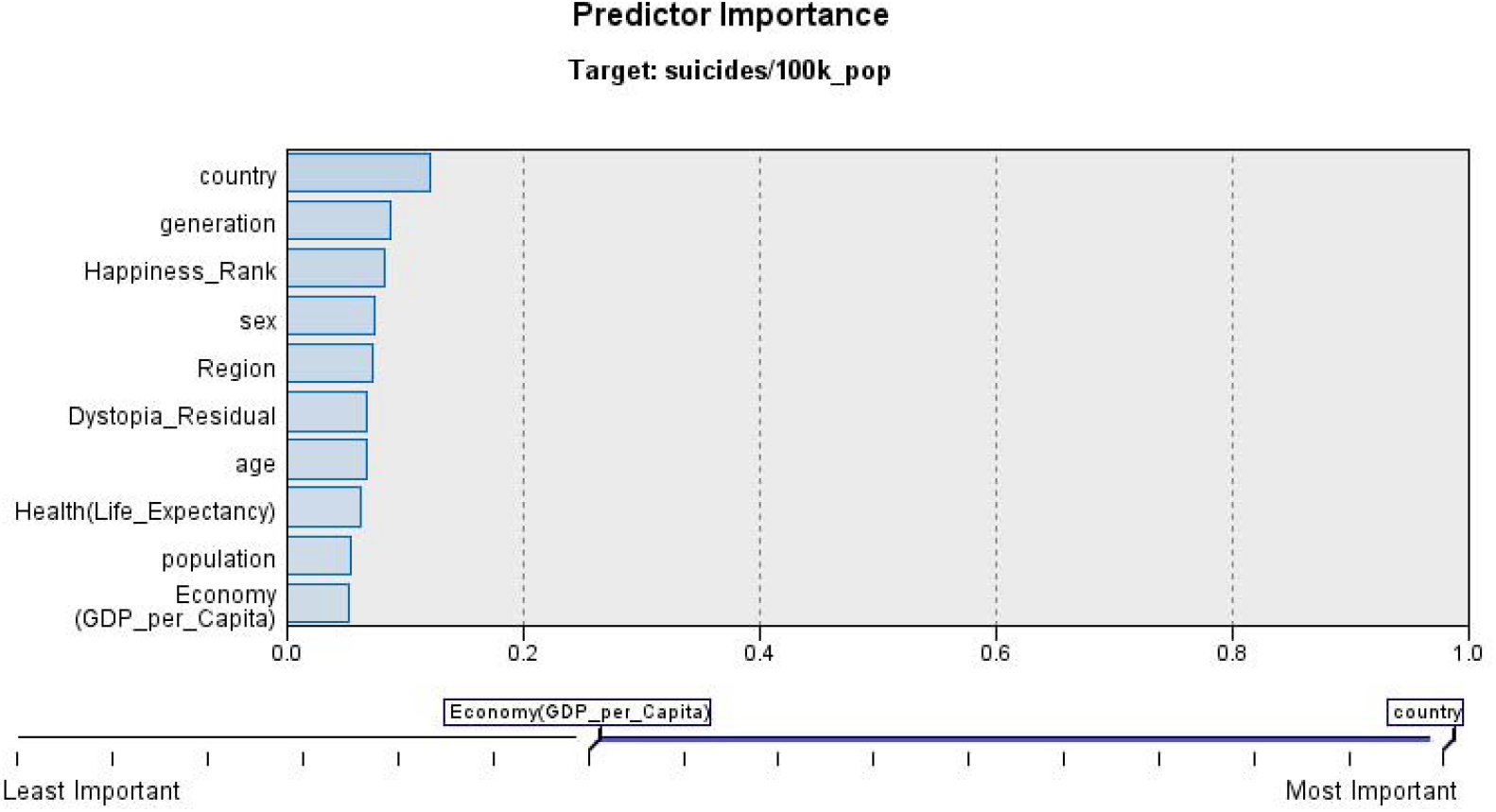
Predictor Importance: Neural Network with 2 hidden layers.

The key predictors are: (a) Country: 12%, (b) Generation: 9%; (c) Happiness rank: 8%; and (d) Gender: 7%

**Fig. 17 (e):**
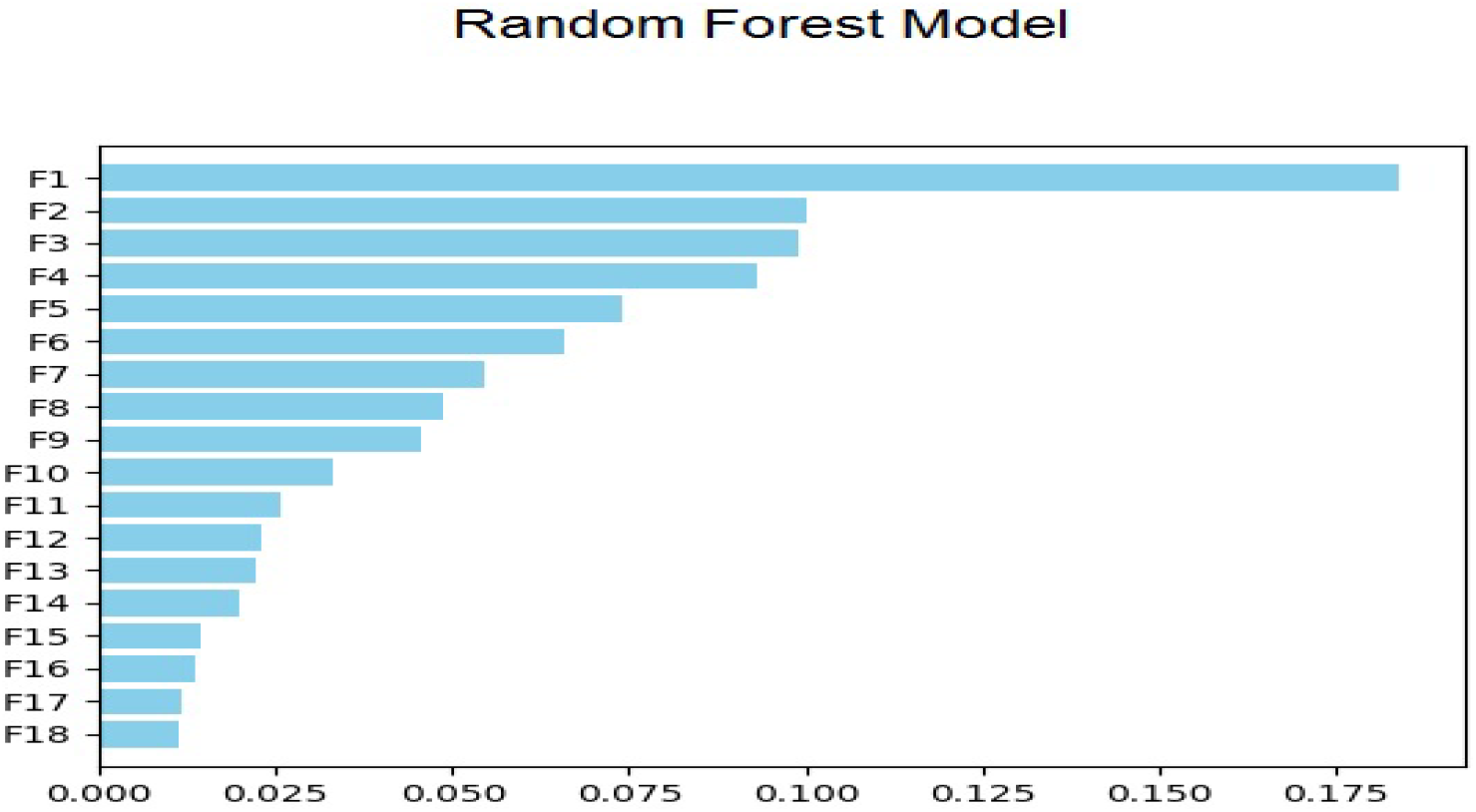
Predictor Importance: Random Forest Model.

The key predictors are: (i) Gender: 17.5%; (ii) Generation; and (iii) Country with percentage not being given for the latter two as these have been interpreted for random forest algorithm’s voting mechanism.

**Fig. 17 (f):**
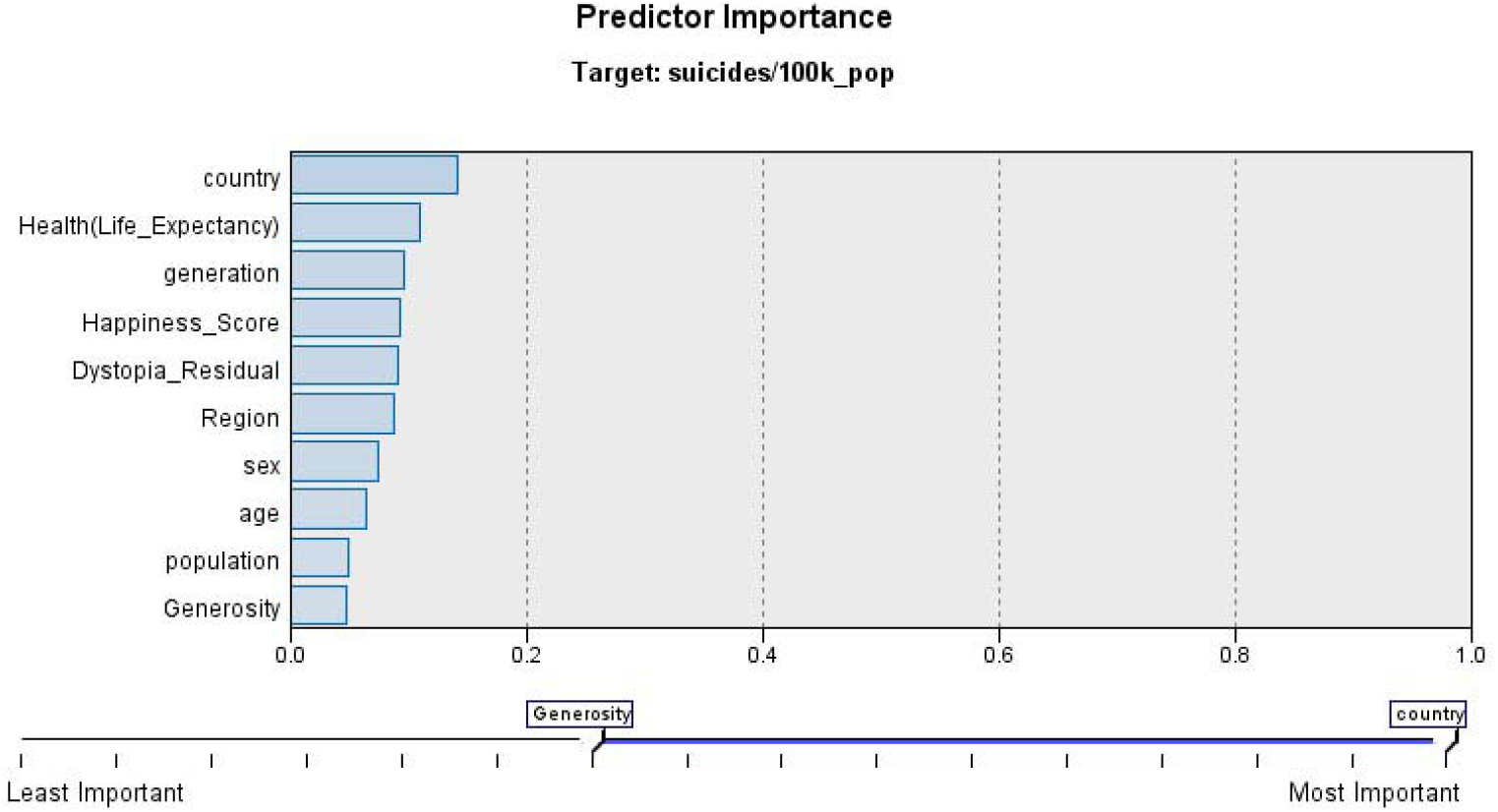
Predictor Importance: Neural Network with 1 hidden layer.

The key predictors are: (i) Country: 14%; (ii) Health: 11%; and (iii) Generation (10%).

This neural network had one hidden layer with 4 nodes in the hidden layer besides the bias unit. Since neural network with one hidden as well as two hidden layers were run for the same dataset, accuracy becomes an acceptable way to decide which findings should be given greater importance. The accuracy of single hidden layer neural network was 85.2% as compared to 90% for a two hidden layer neural network. These details are discussed in later in this study.

**Fig. 17 (f):**
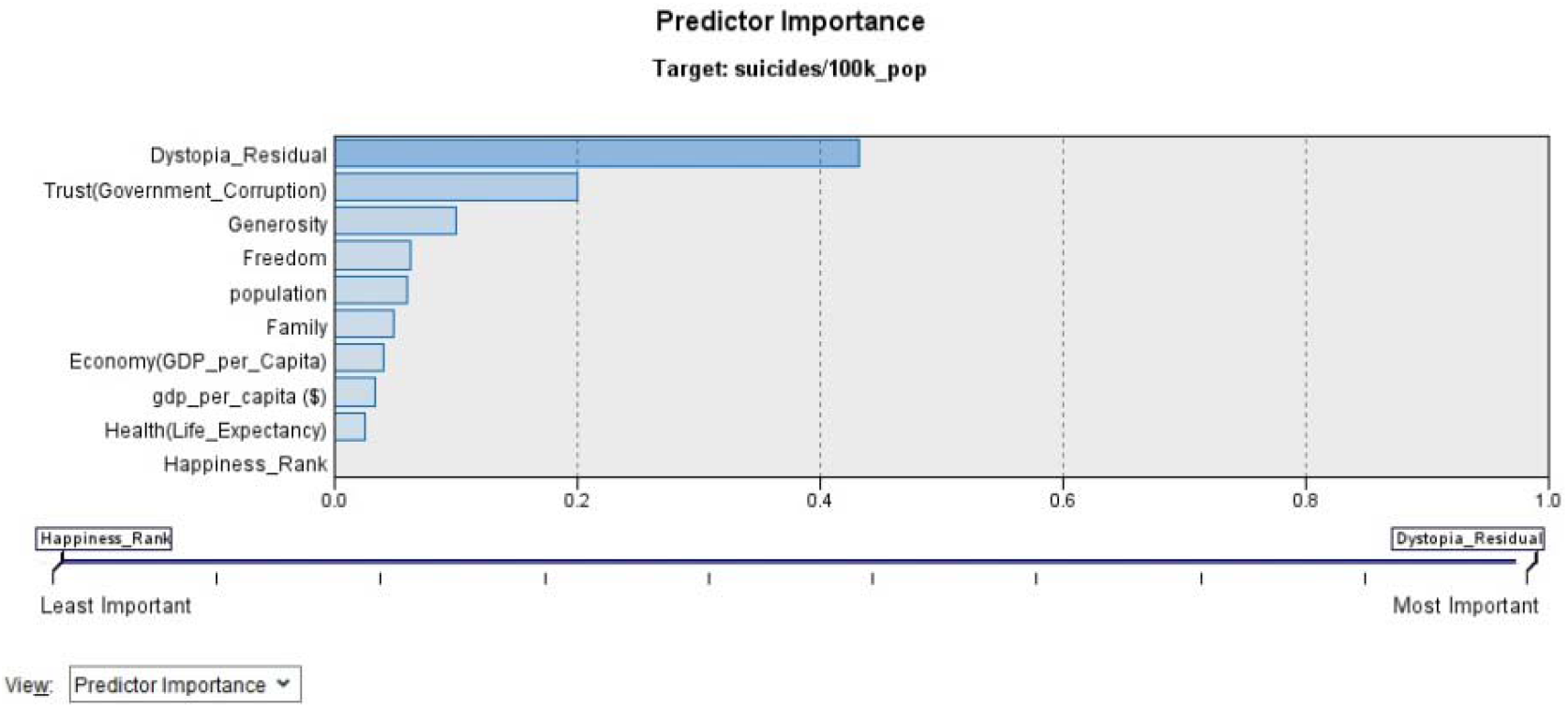
Predictor Importance: Regression Model.

The key predictors are: (i) Dystopia_Residual: 43%; (ii) Trust: 20%; and (iii) Generosity: 10%.

**Fig. 17 (g):**
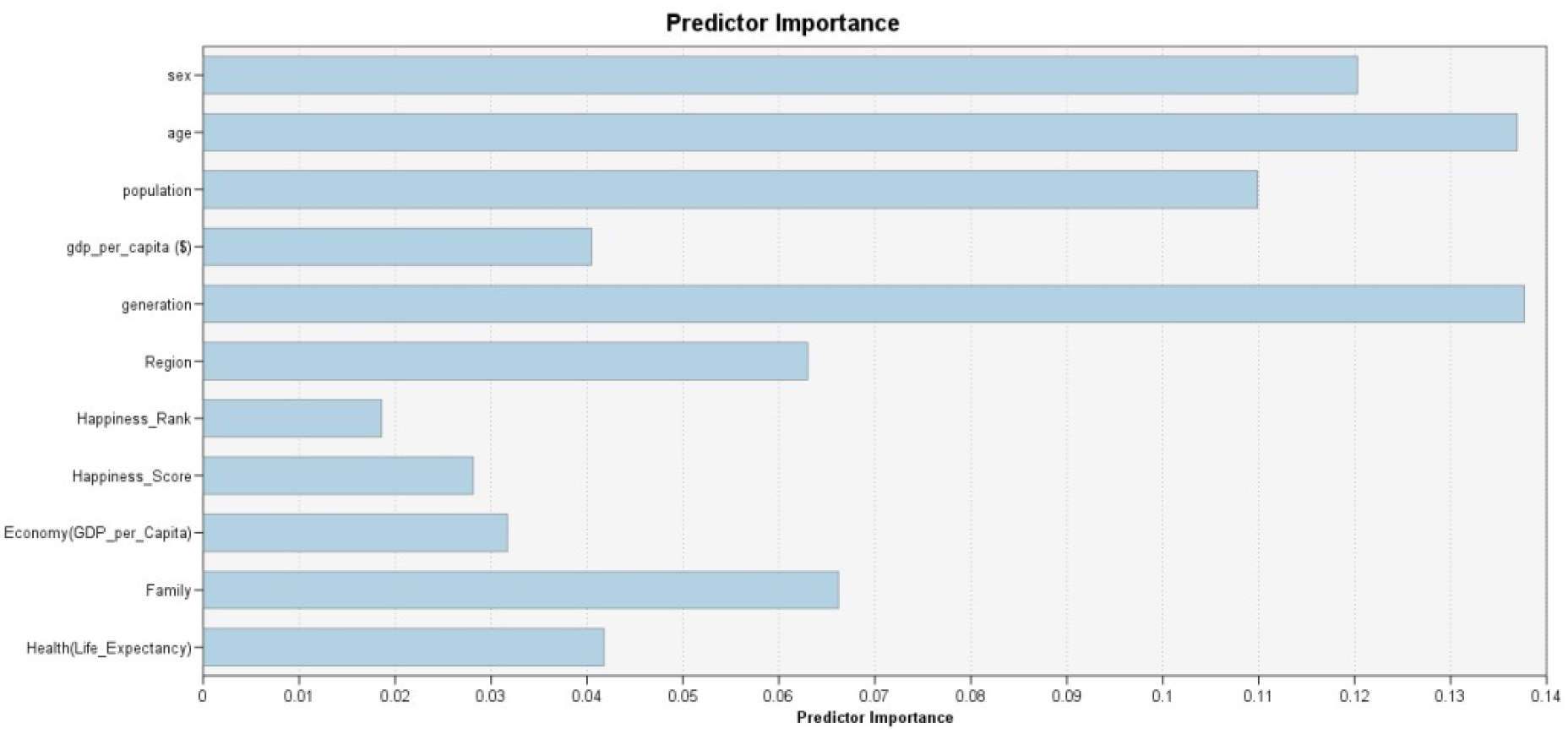
Predictor Importance: Random Trees Model.

The key predictors are: (i) Age: 14%; (ii) Generation: 14%; (iii) Gender: 12%; and (iv) Population: 11%

To enhance the accuracy of prediction, a fully connected neural network with 2 hidden layers was used wherein the first hidden layers had, besides the bias unit, 10 nodes and the second had 8 nodes. A schematic representation of the neural network with 2 hidden layers is given below:

**Fig. 18 (a):**
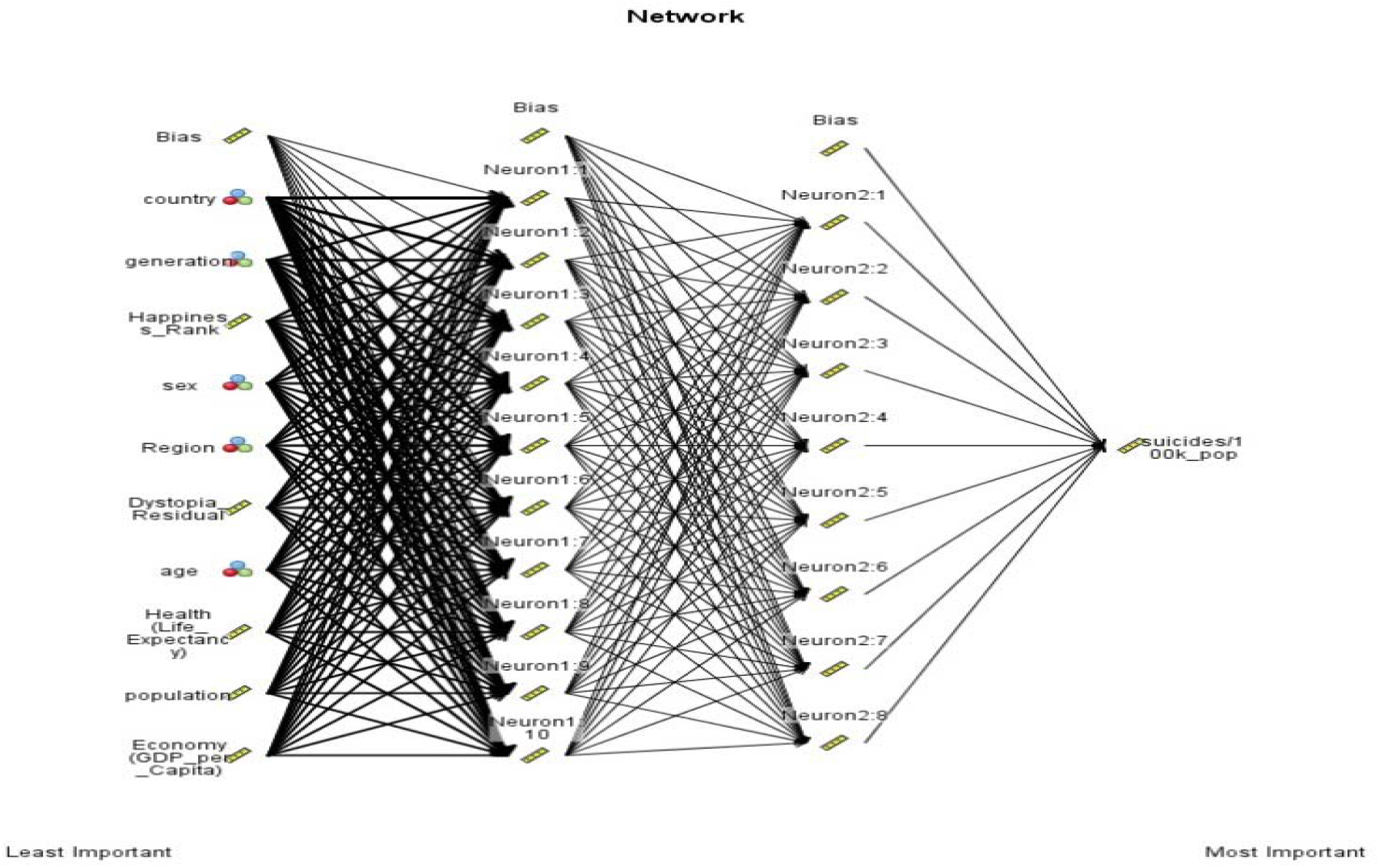
Fully Connected Neural Network with one hidden layer.

The network above provided an accuracy of 90% as shown below:

**Fig. 18 (b):**
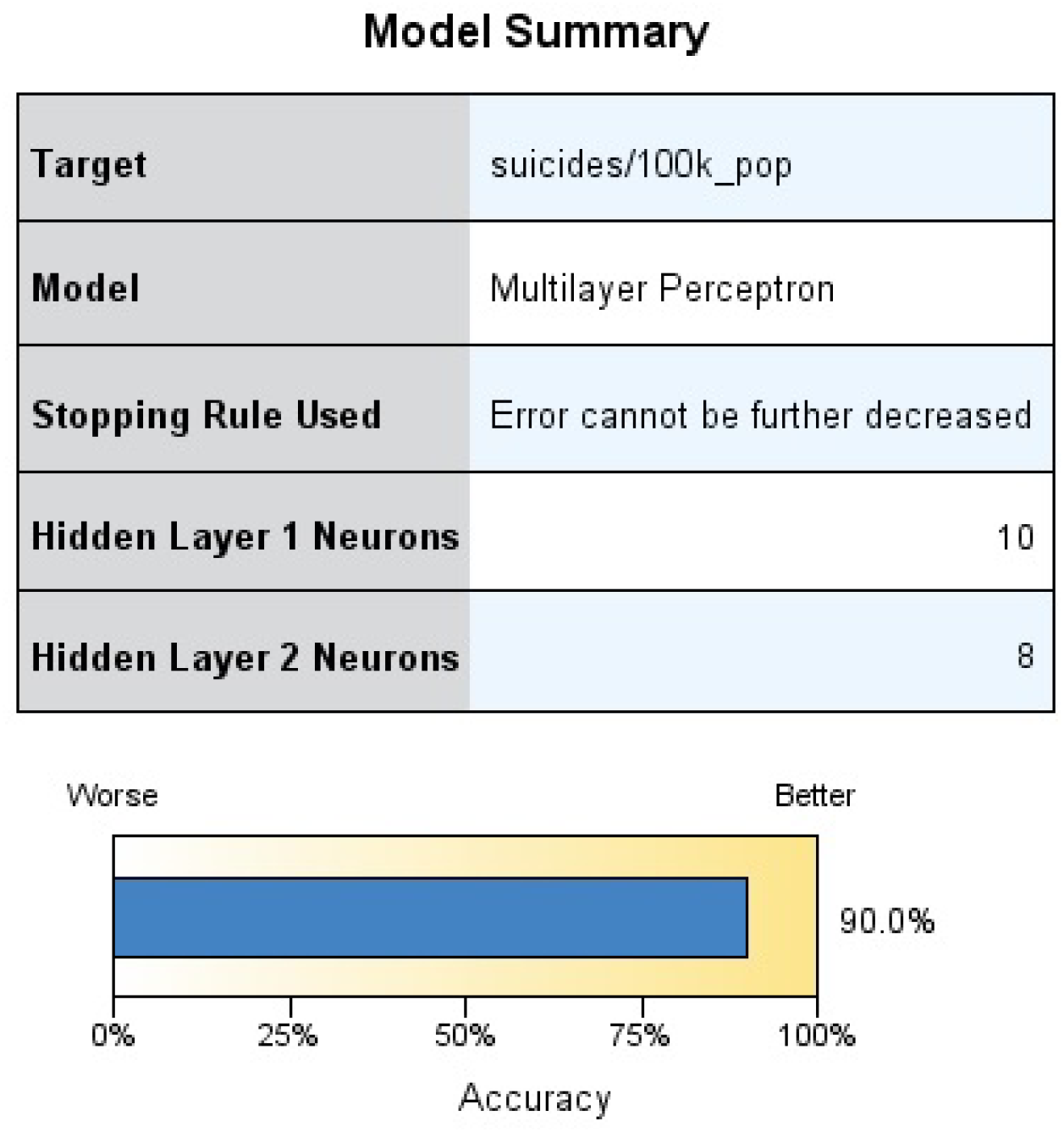
Accuracy of Neural Network with 2 hidden layers.

The data above is summarized as below as regards factors that are the key predictors of suicide:

Assigning weights like done earlier leads to Age being the key predictor followed by Country and then Gender. Adding Country specific data, therefore, leads to Country emerging as the second most important predictor after Age.

## CONCLUSIONS

Summarizing Table 1 and Table 2 leads to the conclusion that ‘Country’, ‘Generation’, ‘Gender’ and ‘Age’ are important predictors of suicide. Since, ‘Generation’ is very closely linked to age groups (wherein the age groups 15-24 and 25-34 are merged together as ‘Millenials’ but other age groups are given a different name) so it would be in order to conclude that the key predictors are Age, Country and Gender with the older male population being the highest risk category. Country and Gender are the other key predictors.

**Table 2:** Identifying the Key Predictors (factors) of Suicide based on Suicide and Happiness data.

| Predictor | Most Important | 2<sup>nd</sup> in importance | 3<sup>rd</sup> in importance |
| --- | --- | --- | --- |
| Age | Gen_Linear<br>Random Trees |  |  |
| Country | Neural Net.: 1 hidden layer<br>Neural Net: 2 hidden layers | Generalized Linear | Random Forest |
| Dystopia_<br>Residual | Regression |  | CHAID |
| Gender | Random Forest | CHAID | Generalized Linear<br>Random Trees |
| Generation | CHAID | Random Forest<br>Random Trees<br>Neural Net: 2 hidden<br>layers | Neural Net.: 1 hidden<br>layer |
| Generosity |  |  | Regression |
| Happiness_<br>Rank |  |  | Neural Net: 2 hidden<br>layers |
| Health |  |  | Neural Net.:1 hidden layer |
| Trust |  | Regression |  |

\* Age here presents a combined view of age group, lower-end and upper-end of age group

The fact that while ‘Country’ emerges as a key predictor but country specific characteristics do not emerge as very important predictor needs further discussion. Since happiness data is scaled for (i) Happiness Score (scaled: 2.9-7.54); (ii) Economy (scaled: 0.09-1.74); (iii) Family (scaled: 0.43-1.61); (iv) Health (scaled: 0.01-0.95); (v) Freedom (scaled: 0.03-0.66); (vi Generosity (scaled: 0-0.84); (vii) Govrnment_trust (scaled: 0-0.46); and (vii) Dystopia_residual (scaled: 0.38-2.9) of the 11 features and the scale is different so some error is likely to creep into the model, especially because the suicide data is not scaled. However, this was not considered to be very serious. A more important source of error came from the fact that happiness reports provide this data for each country and it was assumed to apply equally to all age groups. That may not be very accurate and a more detail study could be conducted where age-wise data and perception on each of these parameters is also taken into account for each age group as suicide is closely linked to perception of the external world. Further, it was felt that increasing happiness report data over a longer duration would help identify more factors that can help predict and, then, prevent suicides.

Since Age, Country and Gender are the key predictors of suicides, effective strategies for suicide prevention can be targeted at the older generation with added focus on older male population. This finding integrates with the recommended approach for suicide prevention by Pan American Health Organization and World Health Organization by identifying the vulnerable groups.

**Fig. 19:**
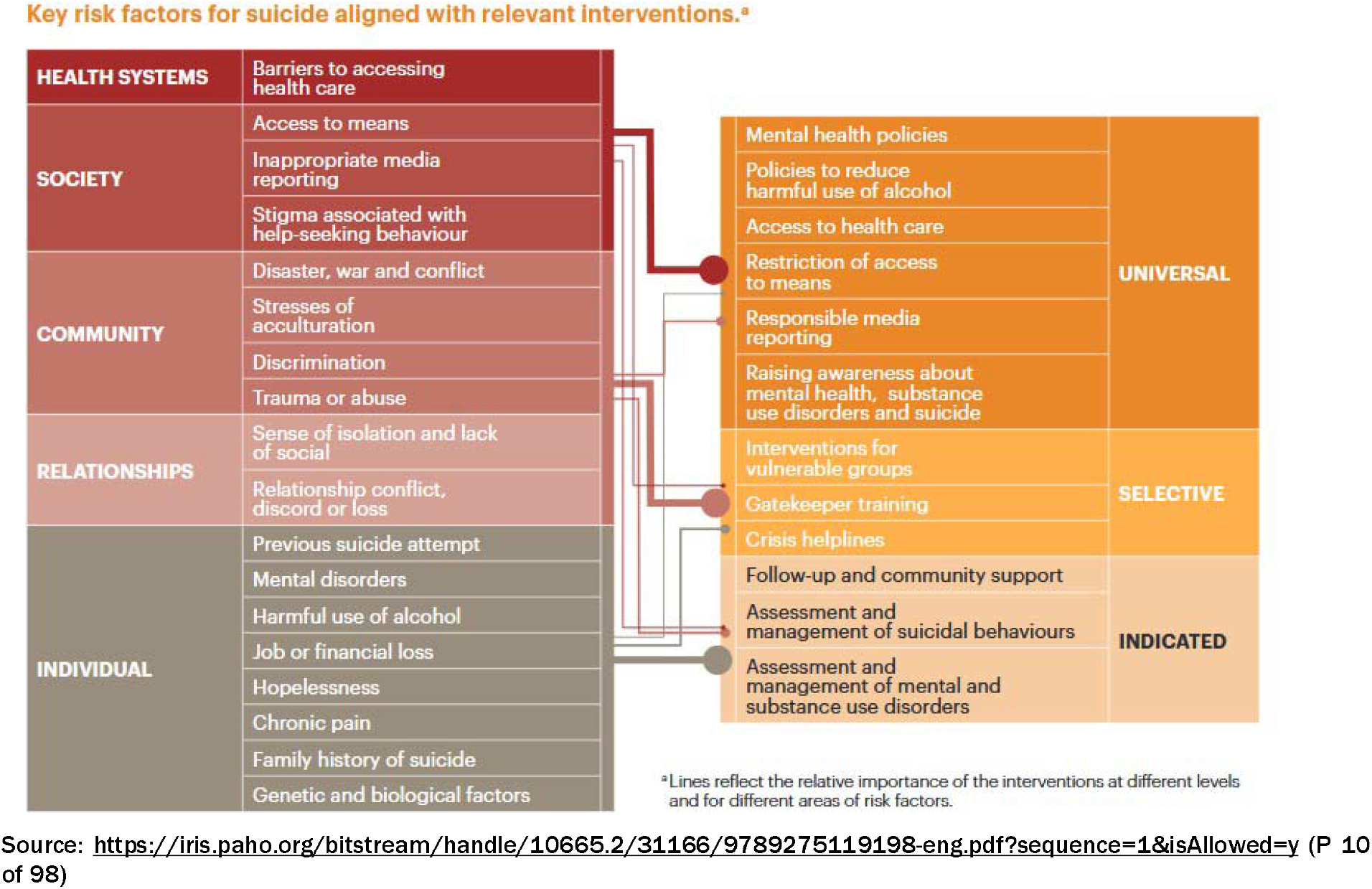
Suicide Prevention Strategies and Interventions

Having identified the high risk category, suicide prevention strategies could, in the short term focus on the vulnerable sections of society i.e. the older generation with males being even more vulnerable than females. Country specific factors are likely to take more time to produce results as these may require bigger changes with more time also needed to reach those who need help.

## OPPORTUNITIES FOR FURTHER STUDY

Given below are areas for improvement and opportunities for further study:

1. Suicide tends to be under reported in the elderly as they have means of overdosing that are difficult to report. This could end up under sampling a significant population.
2. Gender: The stress of Covid has changed the suicide dynamic among working women in Japan. As more women enter the work force, economic & wealth factors/stress may mitigate buffer of gender.
3. Subsidizing workers with unemployment benefits dramatically reduces the risk of death by suicide. Looking at countries unemployment benefit access utilization is a factor to consider for further study.
4. Normalizing youth accessing help mitigates depression in this population. AI studies on youth destigmatization of depression, if correlated with increased access/utilization of mental health, further topic to study.
5. Safely storing guns with trigger locks & safes is a politically neutral approach to mitigate access to lethal means and more politically tenable to implement.
6. AI monitoring of movements of mental illness patients with wearables to predict acute risk factors for suicide: variable heart rate, lack of community ambulation (social isolation), geographic changes in patterns of location-high bridges, pharmacy (drugs), hardware store (poison) is grounds for further study. Present AI wearables (Care Predict) are predicting UTIs and fall risk at home. The same can be applied for suicide in high risk mental health population.

## Data Availability

https://samsat.sums.ac.ir/page-Novin/fa/84/form/pId15008

## Notes

### Competing Interest Statement

The authors have declared no competing interest.

### Funding Statement

No external funding was received

